# Multi-ancestry modeling improves fine-mapping resolution, protein prediction, and discovery for proteome-wide association studies

**DOI:** 10.64898/2026.06.29.26356716

**Authors:** Claudia J. Krueger, Matthew Fischer, Tooba Rizwan, Mira M. Kumar, Siyaa Bhargava, Robert Gerszten, Kent D. Taylor, Michael H. Cho, Jerome I. Rotter, NHLBI TOPMed Consortium, Minoli A. Perera, Xiaowei Hu, Ani Manichaikul, Hae Kyung Im, Heather E. Wheeler

## Abstract

Proteomic predictive models are predominantly trained on cis-acting variants in European-ancestry cohorts, limiting power and predictive accuracy in ancestrally diverse populations. We performed *cis*- and *trans*-protein quantitative trait locus (pQTL) mapping and developed protein-prediction models using whole-genome sequencing (WGS) and plasma protein levels (Olink) across four ancestry groups from the Trans-omics for Precision Medicine (TOPMed) Multi-Ethnic Study of Atherosclerosis (MESA): European (EUR, n=1270), African (AFR, n=675), Hispanic (HIS, n=642), and Chinese (CHN, n=366), and a combined population (ALL, n=2953). African-ancestry samples demonstrated improved fine-mapping resolution relative to cohort size, yielding significantly smaller cis-credible sets than European-ancestry samples, consistent with shorter linkage disequilibrium (LD) blocks and greater allele frequency diversity in African-ancestry populations. For the first time, we benchmarked fine-mapping models SuSiE, SuShiE, MultiSuSiE, and SuSiEx with multi-ancestral cohorts, revealing a precision-recall tradeoff driven by model assumptions. Comparing protein-prediction models, multivariate adaptive shrinkage (MASHR) and ultimate deconvolution in R (UDR) outperformed elastic net (EN) regression, with *trans*-pQTL inclusion and fine-mapping improving prediction performance and proteome-wide association study (PWAS) discovery. Applying our models in PWAS of 10 phenotypes, we discovered 68 protein-phenotype associations in All of Us (AoU) that also replicated in Pan-UK Biobank. MASHR and UDR models identified 60% more protein-phenotype associations than EN. Notably, 32 of these associations were not previously reported in the GWAS Catalog. Overall, our study demonstrates the importance of including multiple ancestries in genomic studies to capture the full spectrum of regulatory variation and improve cross-ancestry generalizability.

## INTRODUCTION

The plasma proteome has long been used as a proxy for protein abundance levels throughout the human body because of its minimally invasive accessibility and its ability to capture protein levels from a broad spectrum of tissues^1^. Recent advances in whole-genome sequencing and antibody-based proximity extension assay (PEA) proteomics technology enable interrogation of the plasma proteome at scale across individuals^2^. However, large-scale proteomic profiling remains cost-prohibitive, leading to the development of computational tools like PrediXcan to perform proteome-wide association studies (PWAS), which help elucidate how SNP-mediated regulation of plasma protein levels gives rise to observed phenotypic variation^3^. While PrediXcan enables the imputation of protein levels in genotyped cohorts, existing models often lack cross-population generalizability due to reliance on European-ancestry training data, which fails to account for the varying linkage disequilibrium (LD) patterns and allele frequencies in global populations ^4–13^. In addition, models often have a restrictive focus on proximal regulators (i.e., *cis*-variants within one megabase (Mb) of the protein-coding gene boundaries), disregarding the distal regulators (*trans*-variants outside one Mb of the protein-coding gene boundaries) that contribute to trait polygenicity^9,13–15^.

To address these limitations, we used Multivariate Adaptive Shrinkage (MASHR) to build multi-ancestry protein prediction models, then compared them to baseline single-ancestry Elastic Net (EN) implementations^16–18^. By jointly modeling across diverse populations, MASHR’s empirical Bayes framework identifies shared and unique patterns across ancestries to refine effect size estimates and mitigate sampling noise^17^. Ultimate Deconvolution in R (UDR) builds on the MASHR framework to more efficiently model the heterogeneous sharing patterns typical of genomic data^18^. We recently used MASHR to improve transcriptome-wide association studies (TWASs) across diverse ancestral populations^15^; however, MASHR has yet to be applied to improve PWAS. We also trained models that include *trans*-variants, which account for a substantial portion of protein variance but are often omitted due to computational complexity^9,13,14,19^. The scale of these models, spanning the breadth of the genome, necessitates strategies to distinguish true functional signals from the surrounding genomic noise.

Statistical fine-mapping jointly analyzes all single-nucleotide polymorphisms (SNPs) within a locus to prioritize candidate causal variants for further study and experimental validation. Fine-mapping models are more accurate in identifying causal SNPs when they are built with and applied to data from multi-ancestral populations, as there is improved power and mapping resolution by leveraging differences in LD and allele frequency patterns across populations^20^. Multiple multi-ancestry fine-mapping models have been developed, each with slightly different prior assumptions and modeling, leaving it unclear which model is most effective. We benchmarked the Sum of Single Effects (SuSiE)^21^ model applied to single-ancestry and meta-ancestry input, in its GPU-compatible TensorQTL^22^ and CPU-based R^21^ implementations, alongside multi-ancestry models Sum of Shared Effects (SuShiE)^23^, MultiSuSiE^24^, and SuSiEx^25^, which jointly modeled the individual ancestral populations, to determine their efficacy in prioritizing causal variants in non-simulated multi-ancestral datasets where direct head-to-head comparison has not been previously performed.

We used WGS and proteomic data from TOPMed MESA, a multi-ancestral dataset (n = 2953) comprised of European (n = 1270), African American (n = 675), Hispanic/Latino (n = 642), and Chinese (n = 366) populations, to train genetic prediction models of protein abundance^26,27^. Using PrediXcan, we evaluated out-of-sample predictive performance in the independent UK Biobank (UKB) cohort by comparing four distinct SNP input strategies (*cis*-baseline, *cis*-fine-mapped, *cis*+*trans*, *cis*+*trans*-fine-mapped), each trained using EN, MASHR, and UDR methodologies^28,29^. We tested whether MASHR and UDR, particularly when integrated with fine-mapped variant prioritization, improve cross-population predictive accuracy. To assess the utility of these models, we compare their PWAS performance with S-PrediXcan using GWAS summary statistics for clinically relevant traits from the All of Us (AoU) Research Program^30^ and Pan-UKB^31^ to identify and replicate protein-trait associations, respectively^30–33^. Our results underscore the utility of multi-ancestry models in fine-mapping and PWAS to uncover likely regulatory mechanisms underlying complex traits.

## METHODS

This study was approved by the Loyola University Chicago institutional review board (Project #2014).

### TOPMed MESA Training Data

The TOPMed program, sponsored by the National Heart, Lung and Blood Institute (NHLBI), leverages WGS, multi-omics data, and clinical phenotypes to advance the understanding of fundamental biological processes that underlie complex conditions such as cardiovascular, respiratory, and sleep disorders^26^. We leveraged WGS data from the TOPMed Freeze 10b (GRCh38) dataset (phd008693.1) for model training^27,34^. Within TOPMed, we analyzed proteomic data from MESA, a community-based cohort study designed to determine the prevalence, determinants, and progression of subclinical cardiovascular disease^27^. MESA recruited men and women aged 45-84 free of clinical cardiovascular disease at baseline from six different locations in the United States and from four major race/ethnicity groups, which included African American (AFR), Chinese (CHN), European (EUR), and Hispanic/Latino (HIS)^27^. Data are available via the dbGaP study NHLBI TOPMed: MESA and MESA Family

AA-CAC (phs001416.v4.p1). WGS and variant calling have been previously described^34^. Protein levels from the TOPMed MESA participants were obtained from blood plasma samples and measured using the antibody-based Olink Explore Assay 3072 proximity extension assay (PEA) platform. The TOPMed MESA study includes participants recruited by six centers across the United States from four self-reported race/ethnic groups: EUR, AFR, HIS, and CHN. WGS and plasma protein data were available for 3311 individuals.

### TOPMed MESA Genomic Data Quality Control

We received variant calls from the TOPMed Freeze 10b WGS dataset, provided by the TOPMed Data Coordinating Center, in VCF file format by chromosome for chromosomes 1 through 22 and X. We converted VCF files to PLINK bed/bim/fam format using bcftools^35^, combined chromosomes, then split by population, resulting in files for each of the four populations, as well as a fifth population of combined ancestries (ALL). For each individual population and the combined population, we removed insertions or deletions (indels) and multiallelic SNPs to accommodate biallelic assumptions of downstream analyses, then excluded SNPs with missing genotype data > 0.02, minor allele frequencies (MAFs) < 0.01, and Hardy-Weinberg Equilibrium (HWE) p < 1e-6 using PLINK v1.9^36^. For the X chromosome, we only applied HWE filtering to variants within the pseudoautosomal regions (PARs), based on GRCh38 positions, and we coded male genotypes as either 0 or 2 within the non-PAR regions, to reflect hemizygosity. We excluded individuals with PLINK v1.9-estimated pairwise relatedness > 0.104, corresponding to second-degree relatives or closer (EUR: 16, AFR: 16, HIS: 36, CHN: 22).

Prior to principal component analysis (PCA), we filtered out SNPs with a genotype missingness rate > 0.01, removed the human leukocyte antigen (HLA) and high LD regions^37^, then LD pruned using PLINK v1.9 with a sliding window of 200 SNPs, advanced by one SNP at a time, removing one SNP from each pair with r² > 0.3. We independently performed PC-AiR analysis from the GENESIS R package to confirm population clustering after filtering^38^. PC-AiR first uses the KING algorithm to estimate pairwise kinship within each population, dividing individuals into unrelated and related subsets prior to PC estimation. During this process, we identified distinct technical clustering artifacts; specifically, a sharp stratification along PC3 isolated a subset of individuals from the main cohorts. To maintain population homogeneity and to remove potential batch effects, we removed these outlier individuals (ALL: 349, EUR: 115, AFR: 83, HIS: 114, CHN: 37). An additional 10 individuals were removed due to missing covariate data (EUR: 1, AFR: 8, HIS: 1). We proceeded with five populations consisting of EUR (n = 1270), AFR (n = 675), HIS (n = 642), CHN (n = 366), and ALL (n = 2953). We re-performed quality control separately on these final population cohorts starting from the pre-quality control PLINK files, applying the same parameters as above; filtering out SNPs with genotype missingness rate > 0.02, MAF < 0.01, or HWE p < 1e-6. Finally, we performed harmonization of the final PLINK bed/bim/fam files with the original VCF file to ensure no reference (REF) or alternative (ALT) allele flipping occurred during quality control (QC), as allele inconsistencies can arise when using PLINK v1.9.

### TOPMed MESA Proteomic Data Quality Control

We received individual-level intensity-normalized protein level measurements from the TOPMed Data Coordinating Center, with up to three time points per individual at Exam 1, Exam 5, and Exam 6 of TOPMed MESA^27,34^. We grouped the proteomic data into five populations consistent with participants’ WGS data, merging based on sample identifications. Of the original 2941 proteins, we removed 41 due to missing protein level data. We removed two proteins, OID30376 and OID31431, with no variance between individuals. To identify potential plate effects, we computed the top 10 PCs of the normalized protein intensity matrix across all samples using ‘prcomp’ in R^39^. We then examined the distributions of PC1 and PC2 across all 103 proteomics plates using kernel density estimates. All plates centered around a common median with no plate-specific shifts in PC1 or PC2 distributions, indicating no batch effects attributable to plate processing order or identity. We adjusted protein levels by the first ten genetic principal components, age at collection, and sex. We also adjusted for race/ethnicity in ALL. We then took the mean of the residuals across time points to represent a single protein level per protein per individual. We proceeded with 2898 unique protein levels per individual.

### UKB pQTL Replication Data

We replicated our TOPMed MESA ancestry-specific fine-mapped pQTLs in 54,219 UK Biobank (UKB) participants^40^, each with 2923 unique proteins, compiled by the UK Biobank Pharma Proteomics Project^29^. We used the UKB study-provided pQTL summary statistics for our replication analyses. Protein levels were measured using the same antibody-based Olink Explore 3072 platform as TOPMed MESA and inverse-rank normalized prior to analysis^29^. In the original study, array-based genotype dosages underwent quality control, retaining only variants with a genotype missingness rate < 0.1, MAF > 0.01, HWE p > 1e-15, minor allele count > 100, genotyping rate > 0.99, and LD pruning (sliding window of 1,000 variants, advanced by 100 variants at a time, removing one variant from each pair with r² > 0.8)^29^. Association testing was performed using linear regression models, with adjustment for age, sex, batch effects, UKB center, UKB genetic array, time between blood sampling and measurement, and the first 20 genetic principal components^29^.

UKB performed pQTL mapping within six populations: European (EUR, n = 33,187), African (AFR, n = 931), Central/South Asian (CSA, n = 920), Middle Eastern (MID, n = 308), East Asian (EAS, n = 262), and American (AMR, n = 97), as well as a combined multi-ancestral population consisting of all participants (META, n = 34,871). We downloaded summary statistics for all plasma proteins with a similar ancestral population within TOPMed MESA (META, EUR, AFR, EAS, and AMR) using Synapse Python Client (syn52355726, syn51365303, syn51365304, syn51365306, and syn51500434, respectively). We also downloaded the combined UKB ancestral population and compared it to the TOPMed MESA combined population, noting the inclusion of the populations CSA and MID.

### UKB Proteome Prediction Testing Data

We tested our proteome prediction models using individual-level WGS and proteomic data from the UKB Research Analysis Platform. Details regarding sampling and data collection have been described previously^28,29^. DRAGEN population level WGS variants, PLINK format [500k release, Data-field 24308] and OLINK 3K PEA Normalized Protein Expression (NPX) proteomics levels [Data-field 30900] were used for this study. In this analysis, we included participants with both WGS and proteomic data for three populations based on self-reported ethnic group categories from UKB: European (British, Irish, or Any Other White Background; n = 49,009), African (Caribbean, African, or Any Other Black Background; n = 1204), and Chinese (Chinese; n = 148). For PrediXcan analysis, we retained the union of SNPs in all prediction models created during TOPMed MESA training.

### TOPMed MESA Protein Quantitative Trait Loci Mapping

We performed pQTL mapping using TensorQTL’s *cis*-nominal argument (‘cis.map_nominal’) to test associations between all *cis*-SNP-protein pairs, *cis*-empirical (‘cis.map_cis’) to estimate beta-approximated p- and q-values based on 10,000 permutations per protein, and *trans* (‘trans.map_trans’) to test associations between all *trans* SNP–protein pairs within each population^22^. For *trans*-fine-mapping, only associations with a p-value < 5e-5 are stored to reduce computational burden. pQTLs are classified as *cis*-pQTLs if they are within one megabase (Mb) upstream of the transcription start site or downstream of the end of the protein-coding region. All others are *trans*-pQTLs. We classified protein-coding gene regions by mapping the Uniprot identifications^41^ assigned to Olink-measured proteins to Ensembl gene identifications and subsequently retrieving the corresponding genomic coordinates from GENCODE (GRCh38), version 47 (Ensembl 113)^42^. We manually annotated proteins whose genomic coordinates are not included in GENCODE by extracting the corresponding mapping information from Ensembl and included them in our analyses. In cases where more than one PEA binds to the same protein, usually targeting different isoforms, we considered each measurement an independent protein. Because OID21327 targets the products of two genes located on different chromosomes, it was treated as two separate proteins throughout our analyses: OID21327 on chromosome 3 and OID21327.1 on chromosome 5. Because TensorQTL’s *cis*-nominal output does not assign a q-value to individual *cis*-SNP-protein pairs, we used the ‘qvalue’ function from the ‘qvalue’ R package^43^ to calculate false-discovery rates (FDRs) for all tested SNP-protein pairs across all chromosomes. *Trans*-SNP-protein pairs’ FDRs were calculated using TensorQTL’s ‘post.calculate_qvalues’ function with a lambda of 0.85. For subsequent analyses, we applied an FDR threshold of 0.05 to classify statistically significant pQTLs.

### Fine-Mapping Analysis

We fine-mapped TOPMed MESA *cis*- and *trans*-pQTLs using the Sum of Single Effects (SuSiE) TensorQTL^22^ and R^21^ implementations, followed by *cis*-pQTL fine-mapping with Sum of Shared Effects (SuShiE)^23^, MultiSuSiE^24^, and SuSiEx^25^, applied to pQTLs with an FDR < 0.05. For *cis*-pQTLs, we filtered the TensorQTL *cis*-empirical output to include only proteins with a q-value < 0.05, and then fine-mapped all SNPs from our post-QC genotype files within each protein’s *cis*-window. For *trans*-pQTLs, we selected all trans SNPs with a q-value < 0.05 (pSNPs) and fine-mapped all SNPs from our post-QC genotype files that were within 1 Mb upstream or downstream of the *trans*-pSNP. Parameters used for each model were selected to most closely mirror the TensorQTL SuSiE fine-mapping implementation (for a detailed description of parameters applied, see Supplementary Note 1).

We then fine-mapped UKB *cis*-pQTLs with SusieR, SuShiE, MultiSuSiE, and SuSiEx summary statistic-compatible implementations with the 1000 Genomes Project (1KG) phase 3 GRCh37 (release file date 2013/05/02) as a reference panel, applying the same parameters as TOPMed MESA fine-mapping where possible (for a detailed description of input LD data and parameters applied, see Supplementary Note 2). For all fine-mapping models, we reported 95% credible sets, meaning each set had a 95% probability of containing the causal SNP.

### Elastic Net Regression

We trained elastic net (EN) regression models separately in each of the five MESA populations: EUR, AFR, HIS, CHN, and ALL^16^. Initially, for each protein, we used individual SNP genotypes within a *cis*-window to predict covariate-adjusted protein levels and considered this our baseline model. Models were trained using the ‘glmnet’ package in R, with a fixed mixing parameter α = 0.5^44^. We employed a nested cross-validation (CV) framework to optimize model parameters and assess performance. Each model was trained and tested across five outer folds, with the optimal regularization parameter (λ) selected via 10-fold inner cross-validation within each fold. Prediction accuracy was quantified using Spearman correlation (ρ) between predicted and observed protein levels in the held-out outer fold.

We did not include all *trans*-SNP genotypes for EN training due to computational constraints. We incorporated fine-mapping data by using posterior inclusion probabilities (PIPs) as penalty factors. For both *cis* and *cis*+*trans* fine-mapped data, we applied a penalty factor of 1-PIP to SNPs within the 95% SuSiE derived credible sets; thus, SNPs with higher PIPs were less penalized and more likely to be retained in the final model. For both methods, *cis* SNPs outside these credible sets were assigned a default penalty factor of 1, matching the baseline regression’s treatment of SNPs without prior functional evidence. As our main goal of building these models is the ability to test as many proteins as possible for association with complex traits in PWAS, we justified this fine-mapping data leakage by testing protein prediction models for replication in an independent cohort (UKB) and comparing their performance in PWAS.

### MASHR and UDR Modeling

We applied multivariate adaptive shrinkage (MASHR) and Ultimate Deconvolution in R (UDR) to estimate protein-level SNP effect sizes by leveraging *cis*- and *cis+trans*-acting pQTLs derived from TensorQTL, first filtered at a nominal FDR significance threshold of q < 0.05 prior to running MASHR and UDR^17,18,22^. This approach allowed for the inclusion of *trans*-effects which were computationally prohibitive in the EN methodology. Both methods jointly model effect sizes across multiple conditions (populations in our study), so we combined SNPs across the four MESA populations (EUR, AFR, HIS, and CHN). For consistency between MASHR and UDR methods, we first computed Z-scores from each SNP’s effect size divided by its standard error. To maximize SNP coverage, we allowed for missing data in one population per SNP. In such cases, we imputed the missing Z-scores derived from effect size and standard error by averaging the corresponding Z-score values from the other three populations.

For the MASHR implementation, we ran the model separately for each protein, treating populations as distinct conditions. We initialized covariance matrices with canonical, PCA, and Extreme Deconvolution covariance matrices^17^. After modeling adjustment, we split weights by population and selected the top 250 SNPs per protein based on the lowest local false sign rate (lfsr) for either *cis* or *cis*+*trans* SNP input types. For fine-mapping models, we instead intersected MASHR-adjusted SNPs with 95% credible sets derived from SuSiE, selecting the SNP with the highest posterior inclusion probability (PIP) within each set, allowing for ties. Final models consisted of four different SNP input strategies: *cis*-baseline, *cis*-fine-mapped (FM), *cis*+*trans*, and *cis*+*trans*-FM.

### UDR Modeling

For UDR, the computed Z-scores were necessary to run truncated eigenvalue deconvolution (TED) algorithms, which require homoscedastic data. We initialized covariance matrices using the “specialized” procedure described by Yang et al. (2024), adapted from Urbut et al. (2019) and described in the “flash_mash” vignette of the mashr R package^17,45^. This “specialized” initialization includes PCA, flash decomposition, and canonical covariance matrices. We used these matrices to initialize the Ultimate Deconvolution Model prior to fitting, applying the Truncated Eigenvalue Decomposition (TED) algorithm with an inverse Wishart (IW) penalty to estimate the prior covariance structure, as recommended in Yang et al. (2024)^18^. We passed the updated covariance structures into the MASHR pipeline to compute posterior summaries. We followed the same procedures as described for MASHR for SNP filtering, including both lfsr-based selection and SuSiE-based fine-mapping, for final models consisting of four different SNP input strategies: *cis*-baseline, *cis*-FM, *cis+trans*, and *cis+trans*-FM models.

### Ensemble Model

We compiled an ensemble prediction model by combining the weights across all machine learning methods and SNP input strategies. We selected the highest accuracy (ρ) model per protein based on out-of-sample testing in the UKB cohort. We refined the selection to consist of the highest sample sizes of the training and testing populations. The training MESA populations consist of EUR, AFR, HIS, and ALL and the testing UKB populations consist of EUR and AFR.

### TOPMed MESA Fine-Mapping Resolution Evaluation

To compare fine-mapping outputs between TOPMed MESA ancestral populations and fine-mapping models, we applied established fine-mapping benchmarks that included the number of credible sets discovered within each protein level phenotype; credible set size, defined as the number of SNPs within each credible set; high-confidence likely causal SNP assignment, defined as the number of SNPs assigned a PIP ≥ 0.9, and maximum PIP per protein, defined as the highest PIP assigned within any credible set for each protein^23–25,46–48^. When comparing multi-ancestral fine-mapping models in TOPMed MESA ALL, we restricted analyses to proteins with FDR < 0.05 in ALL pQTL mapping. To compare credible set count per protein, credible set size, and maximum PIP per protein distributions, we performed Kruskal-Wallis tests separately for *cis*- and *trans*-pQTLs. Statistically significant Kruskal-Wallis results (p < 0.05) were followed by pairwise Dunn’s post-hoc tests, using unadjusted p-values to determine significance.

We computed pairwise Spearman correlations of maximum PIP per protein across models to assess consistency of causal variant prioritization.

### TOPMed MESA-Discovered pQTL and Fine-Mapping Replication

Among proteins tested, 2896 were present in both TOPMed MESA and UKB. We filtered UKB summary statistics to only include SNPs that were significant in TOPMed MESA with a q-value less than 0.05. To evaluate replication of TOPMed MESA-discovered pQTLs within UKB, we used the π_1_ statistic, which is an estimate of the proportion of true positive pSNP replications. To calculate the π_1_ statistic, we applied the q-value function from the ‘qvalue’ R package to the overlapping UKB pSNP p-value distribution to obtain the π_0_ statistic, which is an estimate of the proportion of false-positive pSNP replications^49^. The π_1_ statistic is then calculated by taking (1 - π_0_)^50,51^. We calculated the π_1_ statistic for the replicated *cis*-pSNPs and *trans*-pSNPs of all five TOPMed MESA populations.

We assessed performance of fine-mapping models SusieR, SuShiE, MultiSuSiE, and SuSiEx in TOPMed MESA ALL using UKB fine-mapping results to represent a ground truth. We excluded TensorQTL analysis due to its incompatibility with full summary statistics input. We considered a TOPMed MESA credible set replicated if at least one SNP within the credible set was present in any UKB credible set for the same protein-level phenotype. For each SNP present in the overlapping tested protein level phenotype-variant pairs between TOPMed MESA and UKB, we defined a true positive (TP) as a SNP assigned to a credible set in both TOPMed MESA and UKB, a false positive (FP) as a SNP assigned to a credible set in TOPMed MESA but not in UKB, a false negative (FN) as a SNP assigned to a credible set in UKB but not in TOPMed MESA, and a true negative (TN) as a SNP not assigned to a credible set in either cohort. From these values, we computed precision, recall, and F1 score.

### Evaluation of Model Predictive Performance

To assess the out-of-sample performance of our trained proteome models, we applied the weights derived from the TOPMed MESA cohort to independent genotypes from UKB. We used PrediXcan to generate imputed protein expression levels based on individual UKB genetic data^3^.

We quantified prediction accuracy via Spearman correlation (ρ) between the predicted protein levels and the observed proteomic measurements available in the UKB. Consistent with established thresholds in transcriptomic and proteomic modeling, we considered proteins significantly predicted with a ρ > 0.1 and a nominal significance of p < 0.05^3,13,32^. We quantified model performance by determining the total number of significant protein models and the rho distributions across each population and model-building strategy.

### Proteome-Wide Association Studies in the All of Us Research Program and UKB

To evaluate the utility of the protein predictive models in association studies, we performed PWAS by using S-PrediXcan on GWAS summary statistics available from the All of Us (AoU) Research Program^30,32,33^. S-PrediXcan integrates GWAS summary statistics with our trained proteome weights to estimate the association between predicted protein expression levels and various traits. AoU includes electronic health records compiled from over 800,000 individuals from diverse ancestral backgrounds, including European (EUR), African (AFR), Admixed American (AMR), East Asian (EAS), South Asian (SAS), and Middle Eastern (MID) ancestries^52^. We used the AoU controlled dataset version 8 released on 2025/02/03.

We conducted PWAS to find protein associations across 10 phenotypes, which were selected from phenotypes with the greatest case sample sizes across all analyzed ancestries. These phenotypes include hyperlipidemia, essential hypertension, sleep apnea, type 2 diabetes, asthma, chronic kidney disease, overweight and obesity, osteoarthritis, major depressive disorder, and anemia. Phecodes and GWAS sample sizes for both AoU and Pan-UKB are listed in Table S2.

For each phenotype, we applied S-PrediXcan using TOPMed MESA-derived protein prediction models, trained on ALL, EUR, AFR, and HIS and the GWAS summary statistics generated by the AoU Research Program from EUR, AFR, AMR, and combined (META) populations. We excluded the TOPMed MESA CHN training model population and the AoU EAS testing population from these analyses due to limited sample size. To account for multiple testing across various proteins and model architectures, we applied a conservative Bonferroni-corrected significance threshold of 1.66e-7 (calculated as 0.05/(average number of proteins per model, n = 2098) x (number of models, n = 144))).

We then evaluated replication of statistically significantly predicted protein level-trait associations in AoU using the META Pan-UKB GWAS summary statistics^30^, which encompass diverse ancestries including European, African, and other non-European populations. Pan-UKB GWAS summary statistics were downloaded using ‘wget’ commands from the Pan-UK Biobank phenotype manifest (2023/03/01). Models used and number of proteins in each model for AoU and Pan-UKB are in Table S3 and S4, respectively. We considered an association successfully replicated if the same proteome prediction model (same method, MESA-trained population, and SNP-input strategy), same phenotype, and same direction of effect, yielded a significant protein-trait pair in the Pan-UKB META analysis (mean Bonferroni-adjusted p-value threshold per phenotype: 2.62e-5).

Finally, to assess if the protein-trait association pairs found in our study had been previously reported, we cross-referenced them against the GWAS Catalog (all associations v1.0 downloaded 2026/02/02)^53^. To account for varying phenotypic nomenclature, we employed additional mapping vocabulary and synonymous terms, as phenotype labels may not match 1:1 with GWAS catalog entries (Table S5). For associations with ambiguous or limited matches, we conducted a manual review to confirm that the cataloged traits were clinically and biologically equivalent to our studied phenotypes.

### Colocalization Analysis

From AoU- and Pan-UKB-replicated predicted protein level-trait associations, we performed colocalization analysis with the ‘coloc’ R package^54,55^ between the *cis*-pQTLs of the identified proteins and the corresponding full *cis*-regions in the AoU GWAS summary statistics. We used default priors and considered shared genetic associations if the region had a posterior probability of colocalization (PP.H4) ≥ 0.8.

To visualize colocalization results, we selected a lead SNP present in both the TOPMed MESA *cis*-pQTL window and the corresponding AoU GWAS *cis*-window. For each study, we computed the median p-value per ancestry group (TOPMed MESA: ALL, EUR, AFR, HIS; AoU: META, EUR, AFR, AMR), then took the median of those ancestry-level medians per study, and selected the SNP with the minimum median p-value across the two studies. Lead SNPs are indicated by their respective TOPMed MESA GRCh38 variant identifications. We calculated AoU r^2^ LD measurements using LDlink’s web-based LDproxy^56^ with the following parameters: 1000 Genomes GRCh38 high-coverage reference, lead SNP position (CHR:POS), a ±1 Mb base pair window, and queries run separately within each super-population (ALL, EUR, AFR, AMR).

## RESULTS

### Replication of pQTLs is Best in Populations with Similar Ancestry

We integrated WGS and plasma proteomic data across multiple ancestral populations in TOPMed MESA, AoU, and UKB cohorts to characterize the genetic regulation of protein abundance variation and its influence on complex traits, leveraging multi-ancestry diversity to improve fine-mapping resolution and protein-trait association analyses (Figure 1). After performing *cis*- and *trans*-pQTL mapping within TOPMed MESA, we quantified the number of proteins associated with at least one FDR-significant pQTL SNP (pGenes) and the number of FDR-significant pQTL SNP-protein pairs (pSNPs) for each analysis. Of the 2899 protein levels tested, *cis*-pGene counts ranged from 2323 (CHN) to 2805 (ALL) across ancestry groups, with *cis*-pSNP counts ranging from 136,757 (CHN) to 514,717 (ALL). *Trans*-pGene counts ranged from 591 (ALL) to 1071 (CHN), with *trans*-pSNP counts ranging from 16,673 (AFR) to 50,226 (CHN) (Figure S1). *Cis*-pSNP counts were substantially greater than *trans*-pSNP counts and scaled significantly with sample size (slope = 138.19, R² = 0.817, p = 0.035), while *trans*-pSNP counts did not.

**Figure 1.**
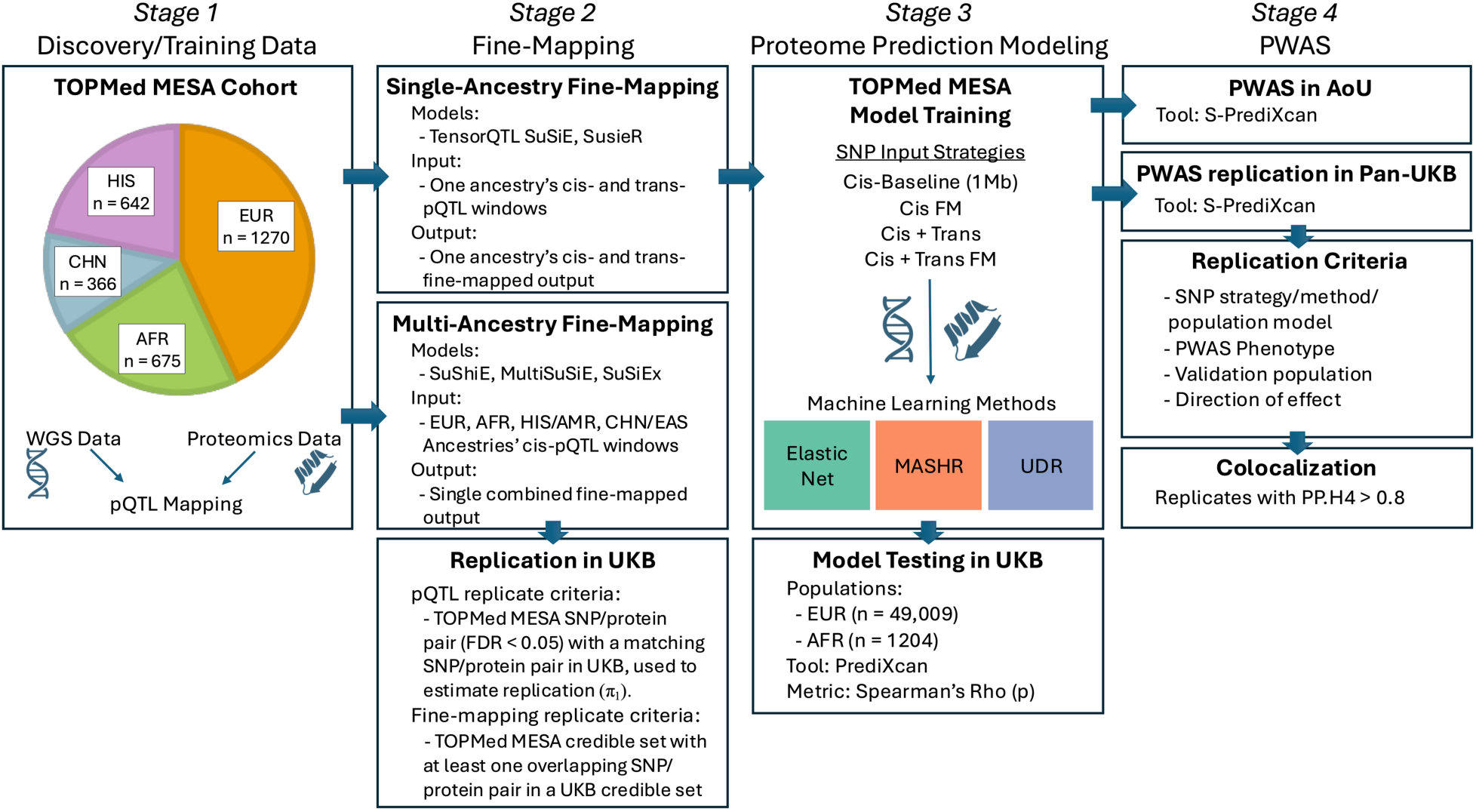
Overall study methodology. (Stage 1) QC of TOPMed MESA cohorts followed by pQTL mapping. (Stage 2) Single- and multi-ancestral fine-mapping within TOPMed MESA followed by replication of pQTLs and fine-mapped outputs in UKB. (Stage 3) Proteome prediction model training within TOPMed MESA followed by out-of-sample testing in UKB. (Stage 4) Proteome-wide association studies (PWASs) using TOPMed MESA-trained models with All of Us (AoU) and Pan-UK Biobank (Pan-UKB) GWAS summary statistics for ten complex trait phenotypes, followed by colocalization analysis. See methods for further details.

To evaluate the replication of pQTLs discovered in TOPMed MESA, we calculated the true positive rate (π_1_) in UKB to quantify replication success. *Cis*-pQTL replication is strongest between similar ancestral backgrounds or larger UKB sample-size populations (META, n = 34,871 and EUR, n = 33,187), with π_1_ ≥ 0.89 for all TOPMed MESA discovery populations (Figure 2). Replication in the UKB African population (n = 931) was moderate depending on training population, with greatest replication from the TOPMed MESA AFR training population (π_1_ = 0.86). Similarly, replication in UKB Chinese (n = 262) was largest from the TOPMed MESA CHN training population (π_1_ = 0.84). The UKB American population was the smallest (n = 97) and had the lowest replication rates (TOPMed MESA HIS to UKB AMR π_1_ = 0.49). TOPMed MESA *trans*-pQTLs had lower replication rates than *cis*-pQTLs in UKB across all populations, particularly those with smaller sample sizes, with most π_1_ values less than 0.75 (Figure 2, S2). Lower replication rates likely reflect reduced statistical power in smaller samples, population differences in genetic architecture between United States- and UK-based cohorts, and the greater noise in *trans*-pQTL discovery.

**Figure 2.**
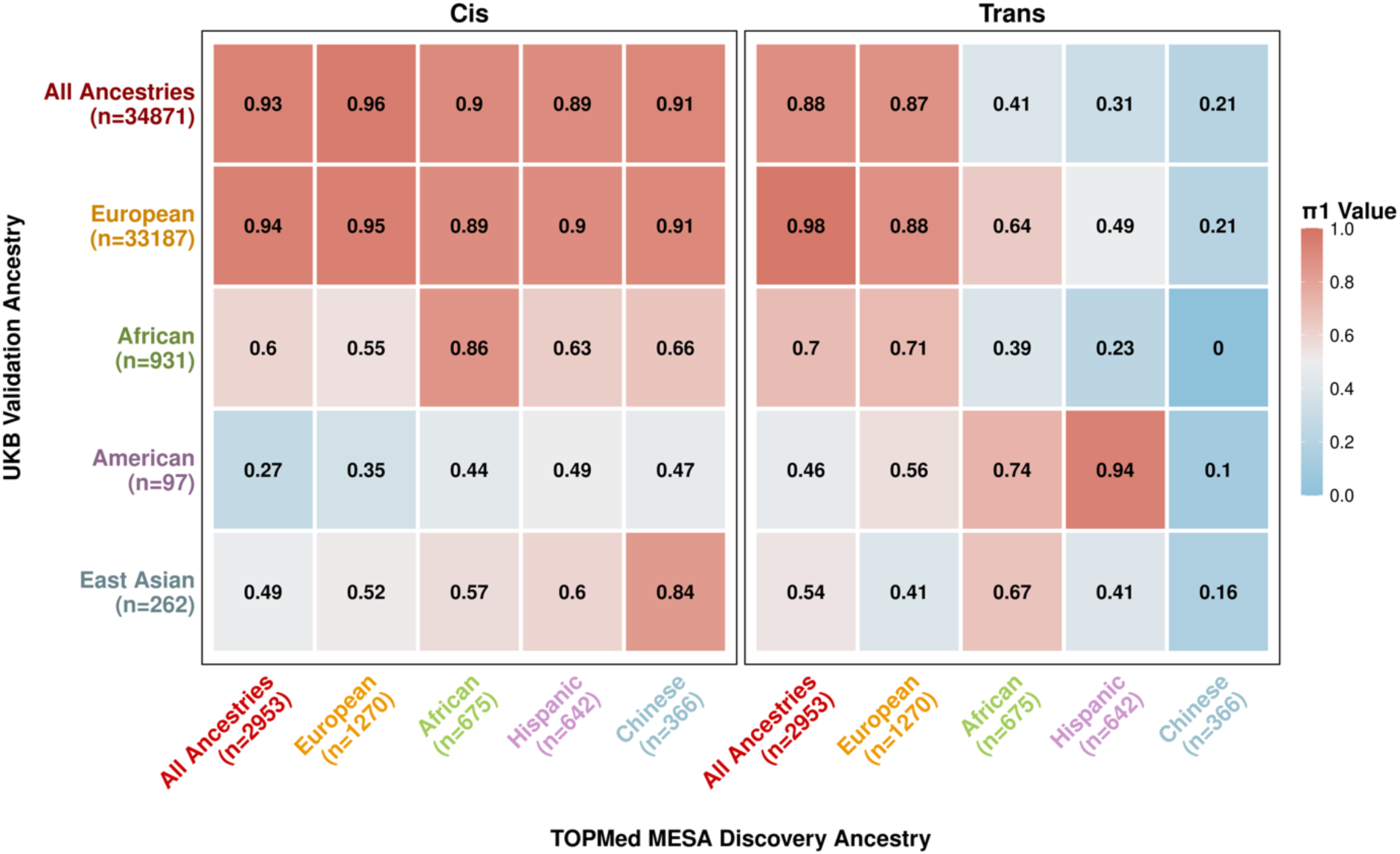
Replication of TOPMed MESA pQTLs in UKB represented by the proportion of true positives (π_1_). Lower replication is colored blue and higher replication is colored red.

### Integration of Diverse Ancestries Improves pQTL Fine-Mapping Resolution

After performing fine-mapping by TensorQTL’s SuSiE implementation within each TOPMed MESA ancestral population, we evaluated the performance by the number of SNPs included in each credible set (Figure 3A) and the maximum PIP within any credible set per protein (Figure 3B). *Cis*-pQTL fine-mapping credible set sizes differed significantly across ancestries (Kruskal-Wallis, p = 8.53e-71), with all pairwise ancestral comparisons significantly different (p ≤ 0.004). ALL produced the smallest credible sets (median = 2, mean = 6.34), followed by AFR (median = 3, mean = 7.95), both outperforming EUR (median = 4, mean = 14.05; ALL vs. EUR: p = 1.94e-50; AFR vs. EUR: p = 1.44e-12). Maximum PIP distributions followed the same pattern (Kruskal-Wallis p = 9.54e-58): ALL and AFR yielded higher maximum PIPs (mean = 0.753 and 0.657, respectively) than EUR (mean = 0.571) (ALL vs.

**Figure 3.**
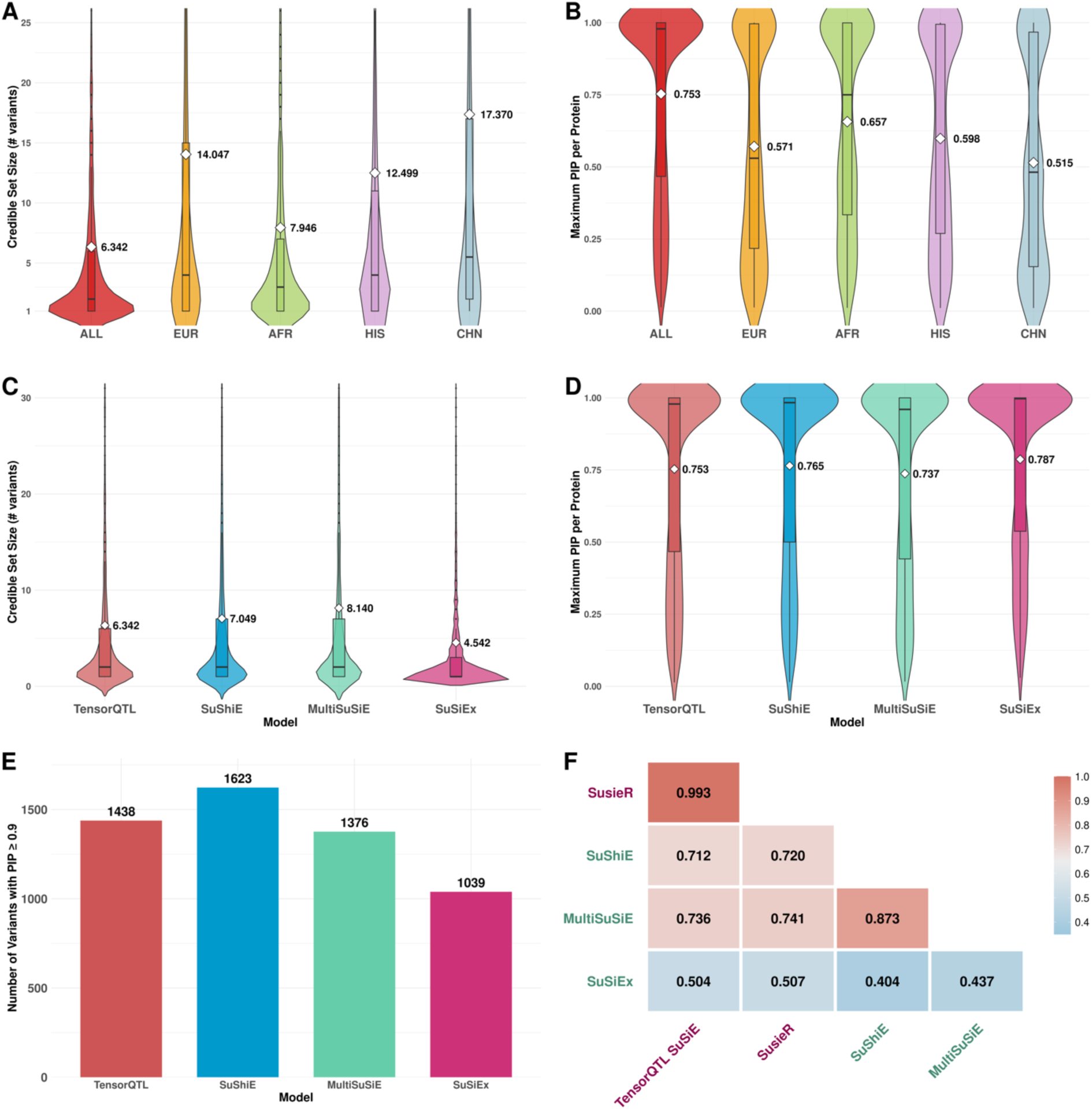
Benchmarking of cis-pQTL Fine-Mapping Models. Violin plots of (A) credible set size and (B) maximum PIP per protein across ancestral populations (ALL, EUR, AFR, HIS, CHN) using TensorQTL SuSiE. For credible set sizes, data points exceeding the y-axis threshold of 30 are present in analyses, but not displayed for clarity. Violin plots of (C) credible set size and (D) maximum PIP per protein across fine-mapping models (TensorQTL, SuShiE, MultiSuSiE, SuSiEx) applied to the ALL population. White diamonds indicate means. Boxplots show median and interquartile range. (E) Count of SNPs assigned a PIP ≥ 0.9, defined as likely causal variants, by each fine-mapping model. (F) Heatmap of pairwise Spearman correlations of maximum PIP assignment within any credible set per protein. Lowest correlations are colored blue and greatest correlations are colored red. Models with single-ancestry input are colored purple, models with multi-ancestry input are colored teal. All pairwise correlations were statistically significant (p ≤ 3.55e-46).

EUR: p = 4.48e-36, AFR vs. EUR: p = 9.60e-6), with all pairwise comparisons significantly different except EUR vs. HIS (p = 0.324) (Figure 3B). These results indicate that ALL and AFR ancestry fine-mapping achieves greater resolution and higher confidence than EUR ancestry alone, consistent with reduced linkage disequilibrium in African-ancestry samples.

Populations with similar genetic ancestries are indicated by similar axis label coloring. Sample sizes are listed below the population label. We calculated the π_1_ statistic from the full p-value distribution of UKB pSNPs that are also present in TOPMed MESA. See Table S9 for UKB SNP input counts.

To assess how diverse ancestral representation within fine-mapping model assumptions influences fine-mapping output, we fine-mapped TOPMed MESA’s ALL population using multi-ancestral models SuShiE, MultiSuSiE, and SuSiEx and compared them to the single-ancestry modeling performed previously with TensorQTL’s SuSiE implementation. Applying single-ancestry models to the ALL population leverages the combined sample size and aggregates LD structure across all TOPMed MESA populations, whereas multi-ancestry models retain population-specific LD to inform causal variant selection (Table S1). Credible set sizes differed significantly across models (Kruskal-Wallis, p = 3.16e-46) (Figure 3C). SuSiEx produced the smallest credible sets (median = 1, mean = 4.54), followed by TensorQTL (median = 2, mean = 6.34), SuShiE (median = 2, mean = 7.05), and MultiSuSiE (median = 2, mean = 8.14), with all pairwise model comparisons significantly different (p ≤ 0.026), except SuShiE vs. MultiSuSiE (p = 0.063). Maximum PIP per protein differed significantly across models (Kruskal-Wallis, p = 1.50e-12). SuSiEx assigned the highest maximum PIPs (median = 0.997, mean = 0.787), differing significantly from all other models (p ≤ 7.38e-7), followed by SuShiE (median = 0.983, mean = 0.765), TensorQTL (median = 0.979, mean = 0.753), then MultiSuSiE (median = 0.960, mean = 0.737). MultiSuSiE assigned significantly lower PIPs than TensorQTL (p = 0.024), and SuShiE did not differ significantly from either (Figure 3D). Using a threshold of 0.9 to classify SNPs as likely to be causal, SuShiE identified the greatest number of high-confidence SNPs (n = 1623), whereas SuSiEx identified the least (n = 1039) (Figure 3E). Maximum PIP within any credible set per protein assignments are more strongly correlated within single-ancestry models and within joint multi-ancestry models than between them, suggesting that shared priors lead to more consistent prioritization of likely causal variants. SuSiEx shows low correlation with other models as it does not borrow strength from a shared prior across ancestries nor pool ancestries jointly, but instead models each ancestry independently, resulting in a distinct set of likely causal variant assignments that differs from both single-ancestry and joint multi-ancestry approaches (Figure 3F).

We also performed multi-ancestry *cis*-fine-mapping with SusieR, SuShiE, MultiSuSiE, and SuSiEx within UKB EUR, AFR, AMR, and EAS populations to evaluate replication of our multi-ancestry TOPMed MESA fine-mapping analyses. Pairwise precision varied across model comparisons (0.105-0.213), with SuSiEx achieving the highest precision against all other models (0.170-0.213), indicating its TOPMed MESA credible sets are most likely to be corroborated by UKB fine-mapping (Figure 4A). SuShiE and MultiSuSiE showed lower precision (0.105-0.137), consistent with their larger credible sets encompassing more candidate variants. Recall showed the inverse pattern (0.038-0.219). SuSiEx had the lowest recall against all UKB models (0.036-0.090), confirming that its stringent credible sets miss a greater proportion of likely causal variants (Figure 4B). Pairwise F1 scores were modest across all comparisons (0.064-0.168), with within-model scores similar across cohorts (0.111-0.126), suggesting comparable self-concordance between TOPMed MESA and UKB regardless of model (Figure 4C). Together, these results reflect a precision-recall tradeoff that mirrors the credible set size and PIP distribution differences observed in Figure 3C-E.

**Figure 4.**
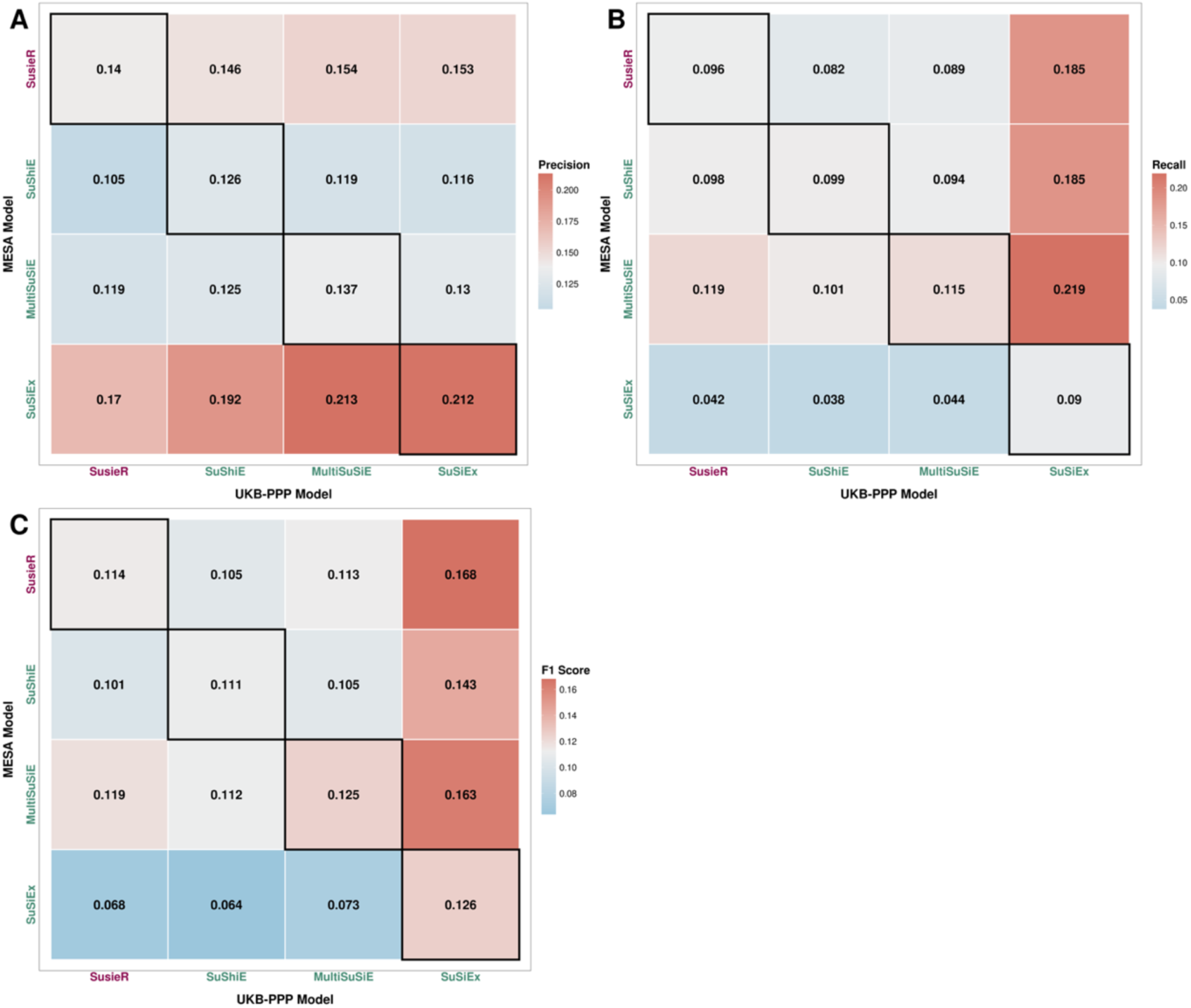
Pairwise fine-mapping model performance metrics. (A) Precision, (B) recall, and (C) F1 scores between fine-mapping models applied to TOPMed MESA (rows) and UKB as “ground truth” (columns). Diagonal cells (black border) indicate same-model comparisons. Color scale ranges from blue (lowest) to red (highest).

### MASHR and UDR Improve Cross-Population Protein Prediction Performance

Before integrating pQTL fine-mapping into our prediction modeling, we first trained *cis*-only protein abundance prediction models in TOPMed MESA EUR and AFR populations using EN, MASHR and UDR (see Methods)^16–18^. We assessed performance by testing the models in the independent UKB cohort. Comparing predicted protein abundance to observed in UKB, we consistently identified a greater number of significant protein models (ρ > 0.1, p < 0.05) with MASHR and UDR than EN in both within-population and cross-population scenarios (Figure 5A). The average significant predictive rate was higher in EN, but fewer models were tested (3091/7470 = 0.4138), compared to MASHR (3475/11,448 = 0.3035) and UDR (3428/10,812 = 0.3171). While EN exhibited a decline in significantly predicted models when applied across ancestries, the generalizability of model performance was improved in the multivariate MASHR and UDR frameworks. Spearman correlation (ρ) distributions confirmed this; EN showed a significant difference in cross-population (AFR train-EUR test median = 0.191; EUR train-AFR test median = 0.164) vs within-population accuracy (EUR train-EUR test median = 0.244; AFR train-AFR test median = 0.223) (Tukey’s HSD, p < 0.001), whereas MASHR and UDR maintained stable ρ distributions across testing populations (AFR train-EUR test median = 0.213 and EUR train-EUR test median = 0.220; EUR train-AFR test median = 0.165 and AFR train-AFR test median = 0.182) (Tukey’s HSD, p > 0.05) (Figure 5B-C). These results demonstrate that while EN performance is dependent on training-population ancestry, multivariate adaptive shrinkage provides a more robust framework for cross-population generalizability.

**Figure 5.**
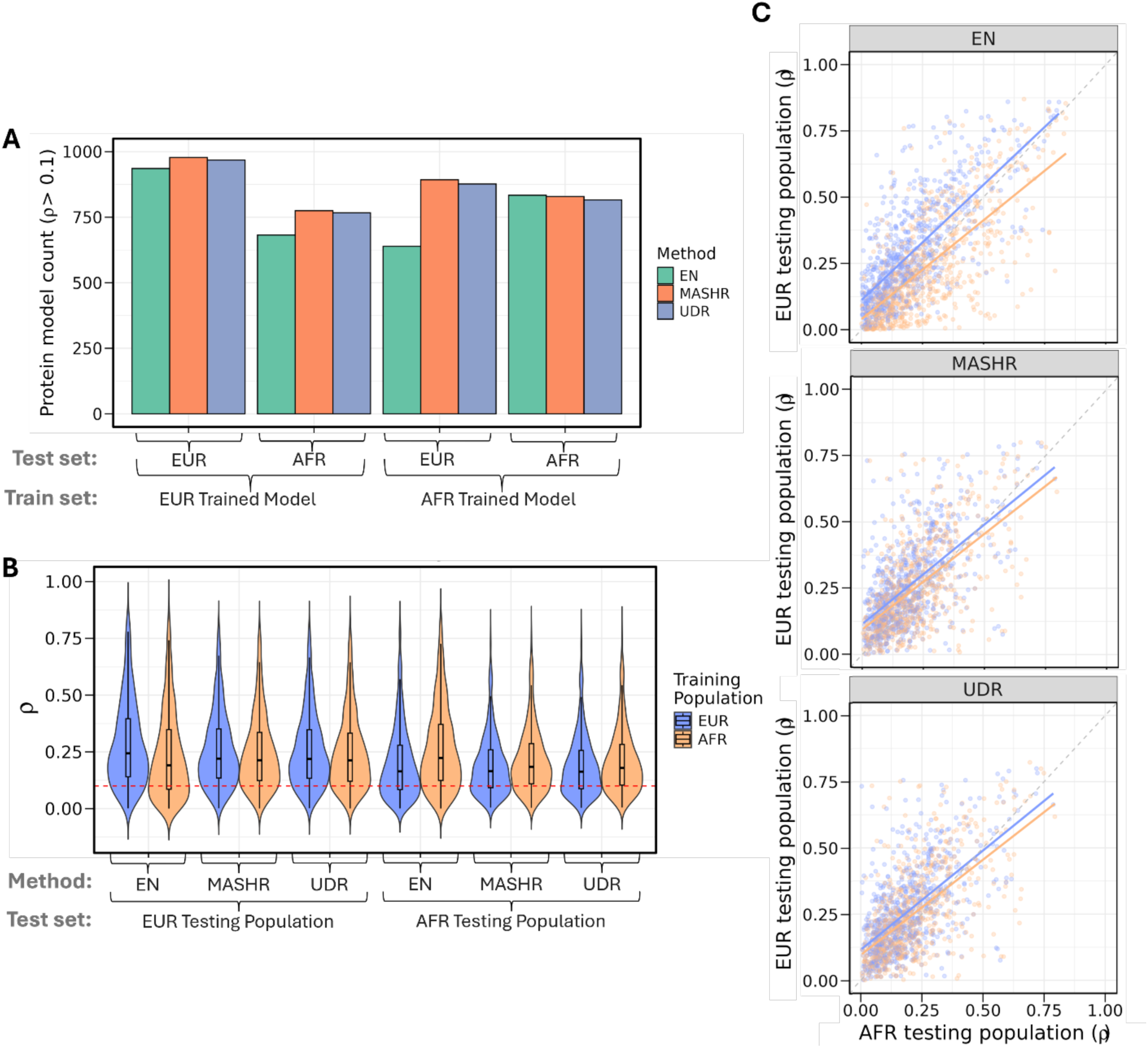
Protein prediction performance in *cis*-baseline TOPMed MESA models. (A) Significant (ρ > 0.1, p < 0.05) protein model counts for each modeling method (EN, MASHR, and UDR). Results are stratified by training ancestry (TOPMed MESA EUR and AFR) and independent testing population (UKB EUR and AFR). Spearman correlation (rho) is calculated between the predicted and observed protein levels. (B) Violin plots display distributions of prediction performance (ρ) across all training and testing scenarios. Analysis reflects a common set of n = 761 with ρ > 0 across all conditions. The dotted red line at ρ = 0.1 denotes the significant performance threshold used for this study. EN models performed worse in cross-population conditions (Tukey’s HSD, p < 0.001), while MASHR and UDR performance did not vary between training-test population pairs (Tukey’s HSD, p > 0.05). (C) Pairwise scatterplot between EN, MASHR, and UDR methods, in which each dot represents a protein (n = 761) comparing EUR (y-axis) and AFR (x-axis) testing population rhos. Blue points and regression lines indicate EUR-trained models; orange points and regression lines indicate AFR-trained models. The grey dashed line represents the identity line (y = x). Statistical significance for the trends observed in 5C is provided by the Tukey’s HSD analysis detailed in 5B.

### Inclusion of *Trans*-pQTLs and Fine-Mapping Improves Overall Predictive Accuracy in MASHR and UDR

We then evaluated four SNP input strategies: *cis*-baseline, *cis*-fine-mapped (FM), *cis*+*trans*, and *cis*+*trans*-FM (see Methods). In EN models, fine-mapping did not significantly alter predictive accuracy (Tukey’s HSD, p > 0.05), likely due to the regularization’s inherent shrinkage of non-informative SNPs (Figure 6A). In contrast, fine-mapped inputs for MASHR and UDR yielded a significant 0.03-0.05 increase in median ρ (Tukey’s HSD, p < 0.0001), effectively matching or exceeding baseline EN performance in within-population contexts. The inclusion of *trans*-pQTLs without fine-mapping in MASHR and UDR may introduce distal noise leading to no performance change compared to cis-baseline; however, *cis*+*trans*-FM models yielded the highest number of significantly predicted proteins across all methods (Figure 6B).

**Figure 6.**
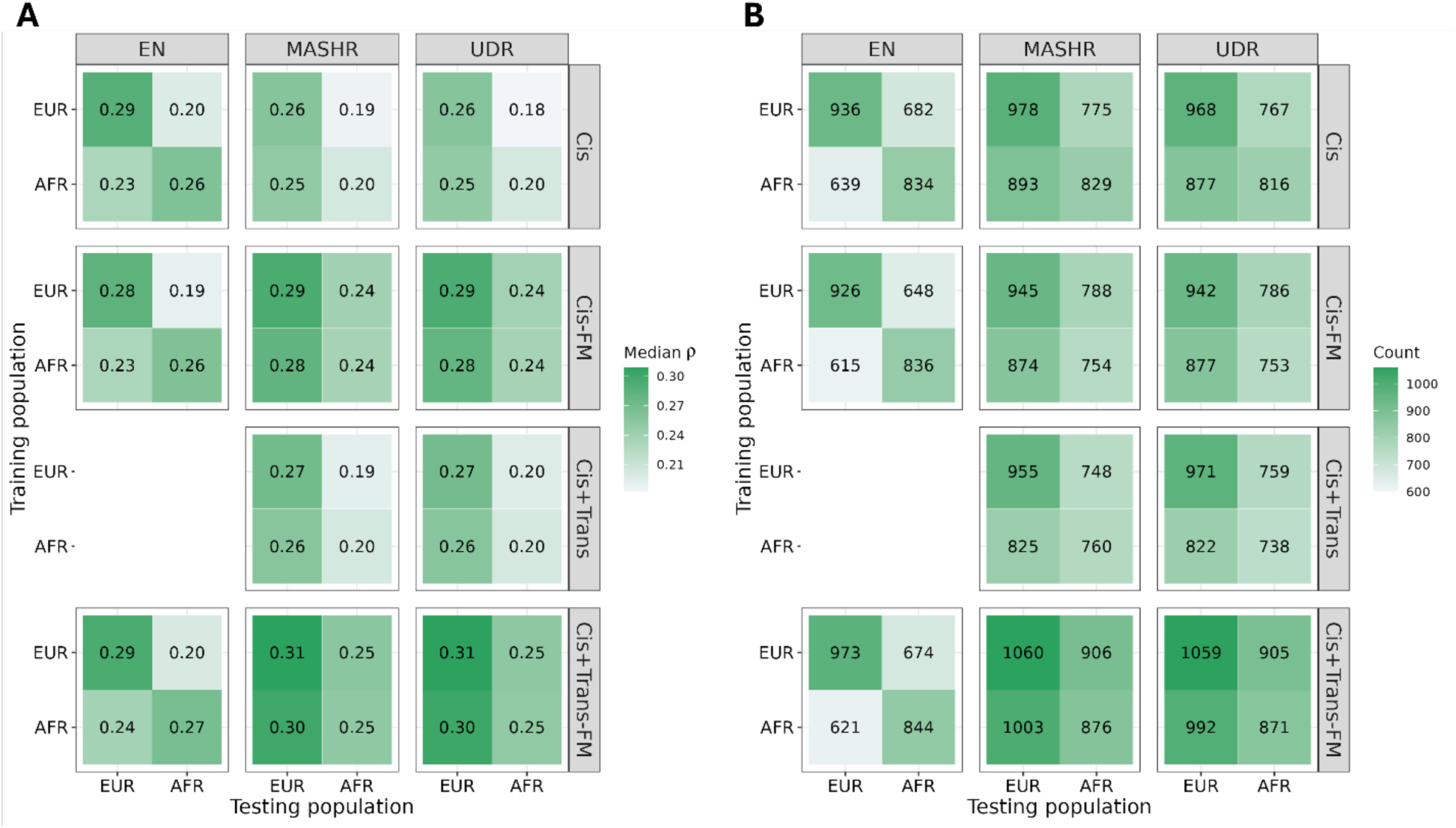
Impact of machine learning methodology and SNP input strategy on predictive performance. Heatmaps displaying (A) the median Spearman correlation (ρ) for a common subset of proteins (n = 532, ρ > 0) and (B) significantly predicted protein counts (ρ > 0.1, p < 0.05) across training ancestries (TOPMed MESA), testing populations (UKB), and modeling frameworks. SNP input strategies include *cis*-baseline (Cis), *cis* fine-mapped (Cis-FM), *cis+trans* (Cis+Trans), and *cis+trans* fine-mapped (Cis+Trans-FM). Cis+Trans modeling was not performed in EN due to computational overhead.

Pairwise comparisons of predictive accuracy showed no significant divergence between MASHR and UDR across all SNP input strategies (Figure S2A). In contrast, overall MASHR (mean = 0.1548) significantly outperformed EN (mean = 0.1454) (Tukey’s HSD, p = 3.69e-29), though performance of individual proteins was heterogeneous across ancestries and SNP input strategies (Figure S2B). While MASHR showed higher means in most EUR-within and cross-population scenarios, AFR-within population testing favored the single-ancestry EN approach, suggesting EN may better capture population-specific architecture in smaller discovery cohorts.

Using MASHR as a representative framework to evaluate SNP input strategies revealed three primary trends (Figure S3A). First, *cis*-fine-mapping (FM) showed that while isolating credible sets removed non-informative noise for some proteins, the strict filtering process likely excluded informative variants for others, leading to specific protein model accuracy declines.

Second, the *cis*+*trans*-FM strategy identified a distinct subset of 19 proteins where accuracy shifted from ρ < 0.25 to ρ > 0.50 (Figure S3C, Table S6). While EN identified similar patterns across SNP input strategies and a subset of 16 proteins improved with *cis+trans*-FM, only 13 overlapped with MASHR, indicating that different modeling algorithms capture distinct distal signals (Figure S3B,C, Table S6). Finally, these results demonstrate that fine-mapping effectively denoises the *trans*-genomic space, allowing models to prioritize high-impact distal regulators that would otherwise be obscured by the high dimensionality of unfiltered *trans*-SNPs.

Given that different modeling methodologies and SNP input strategies captured distinct genetic signals, we sought to maximize our predictive accuracy by developing an Ensemble method. We selected the best model for each protein based on the highest ρ across all UKB training-testing conditions. Of the 2863 protein models in the Ensemble method, 1648 used a MASHR or UDR model and 1215 used an EN model. As expected, the Ensemble method increased predictive accuracy (ρ) (mean ρ = 0.294) compared to the EN *cis*-baseline EUR model (mean ρ = 0.152) (Figure S4), (Tukey’s HSD p < 0.0001). This marked improvement demonstrates that combining strengths across multi-ancestry training, fine-mapping, and *trans*-signals into a combined Ensemble model may better capture the complexities of the human proteome than using a single model across all proteins.

### MASHR and UDR improve PWAS discovery and replication

To evaluate the utility of our developed prediction models, we conducted PWAS using S-PrediXcan across 10 phenotypes using the GWAS summary statistics from AoU, with subsequent replication in the GWAS summary statistics from the META population in Pan-UKB (see Methods)^29–31^. Multivariate frameworks significantly outperformed EN, with MASHR and UDR identifying 96 and 114 total statistically significant associations in AoU (p < 1.66e-7), with 53 and 48 successfully replicating in Pan-UKB (p < 2.62e-5), respectively (Figure 7A, Table S7). In contrast, EN identified 58 associations, with a lower replication count of 31. Binomial general linear model (GLM) analysis of total significant associations vs. total tests confirmed that MASHR and UDR achieved a significantly higher proportion of significant AoU associations per test (0.046% and 0.050%, respectively) compared to EN (0.033%; p < 1.69e-8), indicating that the multivariate advantage persists even when accounting for differences in the number of tested models. Notably, EN discoveries were predominantly limited to hyperlipidemia, while MASHR and UDR captured associations across all ten phenotypes, with replicated associations in seven phenotypes, demonstrating a broader reach into complex phenotypes (Figure 7B). While *cis*-baseline models were the most robust across the varied traits, hyperlipidemia had the most replicated associations (24) only with the inclusion of fine-mapped *trans*-pQTLs (Figure S5A). Analysis by training population revealed that AFR-trained MASHR and UDR models achieved the highest discovery and replication rates across all GWAS populations (AFR, AMR, EUR, and META) (Figure S5B). This confirms that multivariate models successfully share population patterns from high genetic diversity and smaller LD blocks within African-ancestry genomes to build more generalizable weights for PWAS.

**Figure 7.**
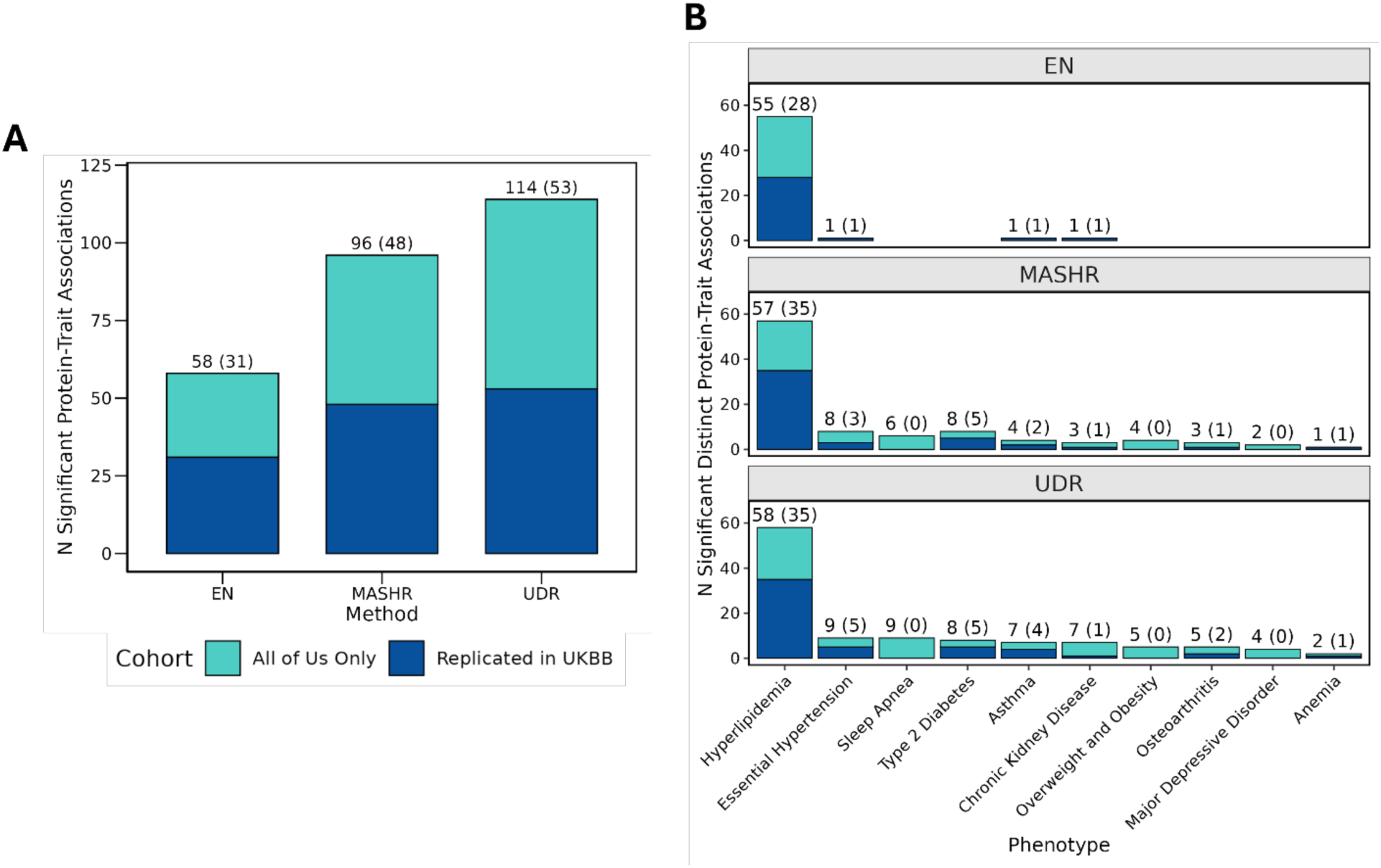
PWAS discoveries in All of Us and replication in UKB, by method and phenotypes. (A) Distinct protein-trait associations per method. Overall comparison of distinct protein-trait association counts between EN, MASHR, and UDR, includes combined results from all models made with each method. Total AoU discoveries (Bonferroni-adjusted p-value < 1.66e-7) are indicated by the height of the stacked bar, with the subset of UKB-replicated associations (Bonferroni-adjusted p-value < 2.62e-5) highlighted in navy. (B) Distinct protein-trait associations by method and phenotype. Overall comparison of distinct protein-trait associations between phenotypes and methods: EN, MASHR, and UDR, includes combined results from all models made with each method. Total AoU discoveries are indicated by the height of the stacked bar, with the subset of UKB-replicated associations highlighted in navy.

We then performed colocalization analysis using all SNPs within the TOPMed MESA *cis*-pQTL genomic window and the corresponding window from the AoU GWAS for each unique protein, TOPMed MESA population, AoU population, training method, and SNP type. 242 of the 270 pairs showed evidence of colocalization (PP.H4 > 0.8) (Table S7), of which there are 11 unique protein-phenotype pairs (Table 1). These results support a shared genetic architecture where SNPs contributing to disease risk may be acting through protein regulation. Of the three protein-phenotype pairs found with MASHR and/or UDR, but not EN, the training population was African, highlighting how the representation of African shorter LD within modeling improves discovery.

**Table 1.**
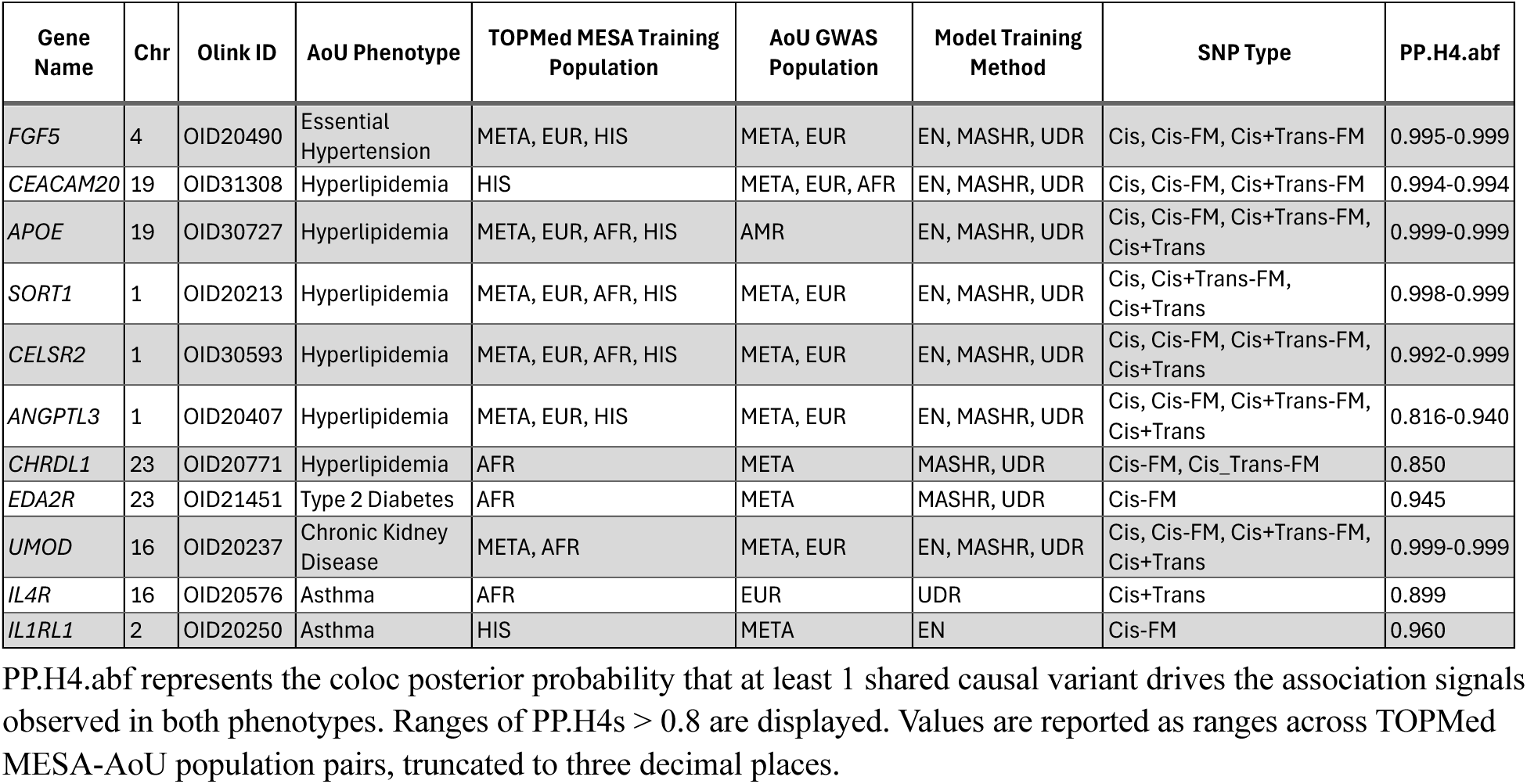
Unique PWAS replicated protein-phenotype pairs with high colocalization probability (PP.H4 > 0.8).

### Identification of Potentially Novel Protein-Trait Associations

Of the distinct 68 replicated protein-trait associations across all models, 32 (47.1%) were not previously reported in the GWAS catalog (Table S8). MASHR and UDR identified the most potentially novel associations compared to EN. While EN identified novel associations exclusively in hyperlipidemia, MASHR and UDR uncovered novel signals in six of the ten phenotypes (Figure 8A). Multivariate discoveries included a link between increased predicted PTH1R levels and essential hypertension risk, and an association between decreased ANGPTL2 abundance and hypertension risk (identified uniquely by UDR). MASHR uniquely identified six novel associations, such as CRLF1 with type 2 diabetes, whereas, UDR and EN uniquely identified 3 and 2 novel associations, respectively (Figure 8B). Furthermore, different ancestry-trained models within the multivariate frameworks contributed unique discoveries; for example, MASHR-AFR identified six associations undetected by EUR or HIS models (Figure S5C).

**Figure 8.**
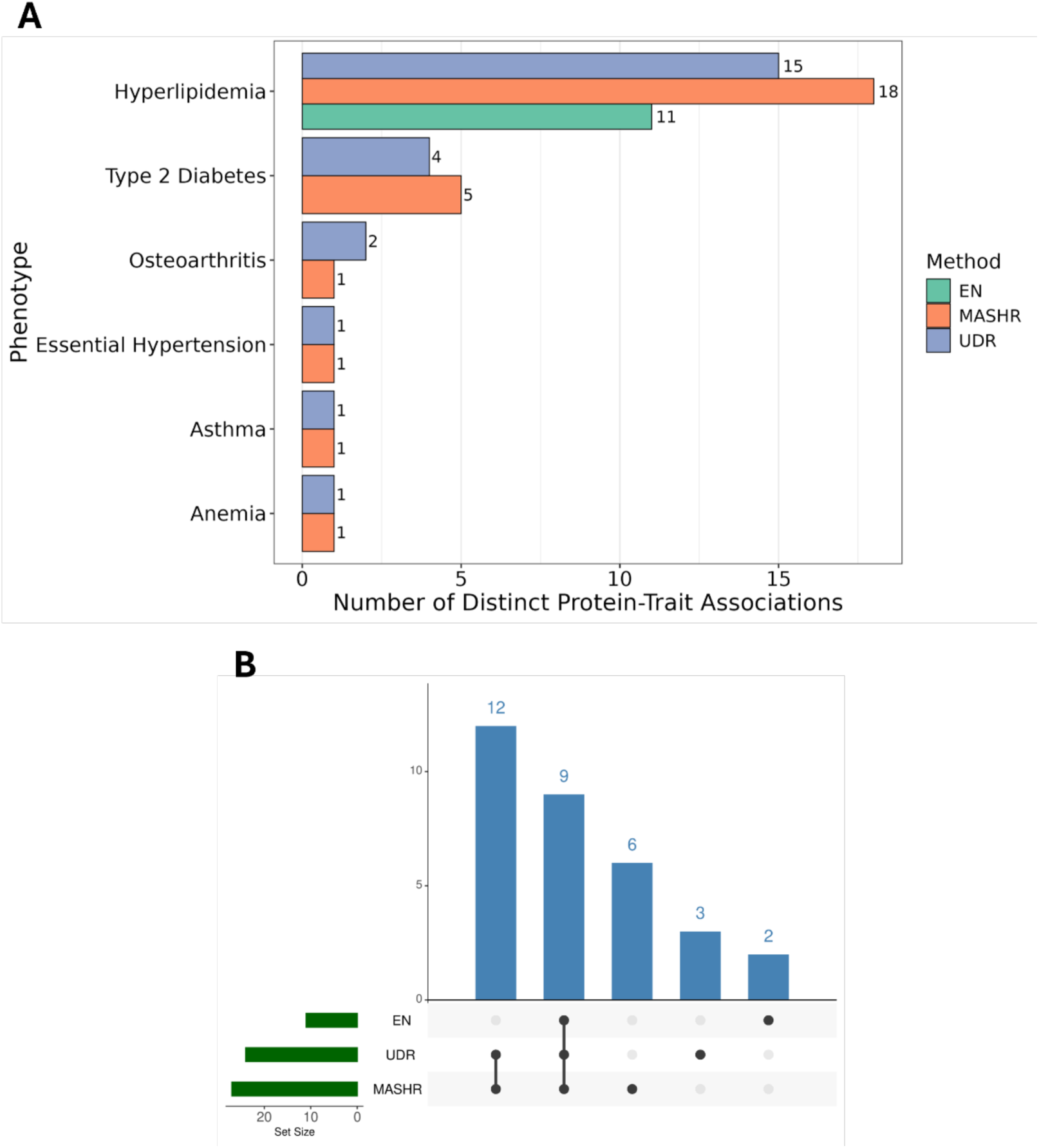
Potentially novel (not in the GWAS catalog) PWAS discoveries across phenotypes and methods. (A) Comparison of significant distinct novel protein-trait associations between phenotypes and methods. (B) Upset plot comparing novel protein-trait associations between models.

## DISCUSSION

We sought to address the persistent ancestral disparities in genomic and proteomic research by developing, benchmarking, and replicating multi-ancestry fine-mapping and proteome prediction for PWAS. Across multi-ancestry fine-mapping models, we observed a precision-recall tradeoff in which models producing smaller, more stringent credible sets achieve higher precision at the cost of recall, offering investigators flexibility to prioritize resolution or coverage while enabling greater overall discovery than single-ancestry models. We also find that multivariate models like MASHR and UDR successfully identify shared and unique patterns of genetic effects on protein abundance across ancestries, mitigating the lack of cross-population generalizability of model performance typically observed in European-centric models.

TensorQTL *cis*- and *trans*-fine-mapping enabled computationally efficient, functionally supported integration of both local and distal regulatory signals across ancestry-stratified models. A shift toward multi-ancestry, *trans*-inclusive modeling provides a robust framework for expanding the reach of PWAS into complex genetic architectures.

*Cis*-pQTLs discovered in TOPMed MESA replicated well within UKB, providing support for the robustness and transferability of *cis*-associations discovered in this study across independent cohorts (Figure 2). On the contrary, TOPMed MESA-discovered *trans*-pQTLs replicated poorly in UKB, particularly in the smaller non-meta and non-European ancestral populations. This reduced replication supports that *trans*-pQTL fine-mapping results may be more susceptible to signal inflation and reduced robustness^57–59^.

TOPMed MESA ALL and AFR ancestries produced the smallest *cis*-credible sets and highest maximum PIPs per protein, outperforming EUR despite AFR’s smaller sample size (Figure 3). These patterns are consistent with the greater haplotype diversity and reduced long-range LD of African-ancestry populations, which enables more precise exclusion of non-causal variants^20,60^. These findings underscore that while sample size contributes to fine-mapping power, ancestral diversity independently drives resolution, and support the prioritization of African-ancestry individuals in multi-ancestry proteogenomic studies to maximize causal variant localization^61–63^.

Multi-ancestry fine-mapping models further improved causal variant resolution by explicitly modeling ancestry-specific LD structure, extending the gains already observed when aggregating diverse ancestries in the ALL cohort over EUR-only approaches. The strong concordance between TensorQTL SuSiE and SusieR (R = 0.993) confirms that their shared statistical framework drives agreement independent of hardware implementation, with TensorQTL offering a computationally efficient alternative^22^. Similarly, SuShiE^23^ and MultiSuSiE^24^ showed high agreement (R = 0.873), reflecting their shared multivariate effect size prior, though SuShiE’s learned pairwise effect correlations modestly outperformed MultiSuSiE’s fixed global sharing structure. SuSiEx^25^ diverged most substantially from all other models, consistent with its assumption of population-independent Bayes factors rather than shared or correlated effects across ancestries.

These modeling differences directly shaped the observed precision-recall tradeoff.

SuSiEx produced the smallest, highest-confidence credible sets but identified the fewest likely causal variants, while SuShiE and MultiSuSiE yielded broader credible sets with greater recall. This tradeoff was confirmed in UKB replication (Figure 4), where SuSiEx achieved the highest precision (0.170-0.213) at the cost of recall (0.038-0.090), and SuShiE and MultiSuSiE maintained higher recall (0.094-0.219) with lower precision (0.105-0.137). Comparable within-model reproducibility across cohorts indicates these differences reflect stable modeling assumptions rather than inconsistent performance.

No single model is universally optimal, though multi-ancestry models should be prioritized when ancestrally diverse data are available. SuSiEx is preferred where high-confidence variant localization is the goal and SuShiE or MultiSuSiE where broader discovery or colocalization is prioritized. Variation in replication across loci further suggests that some genomic regions exhibit greater cross-ancestry shared architecture than others, which may additionally inform method selection. Therefore, characterizing the extent of cross-ancestry shared genetic architecture at individual loci remains an important direction for improving fine-mapping performance, and may ultimately motivate region-specific model selection.

The evaluation of protein expression predictive accuracy across our models demonstrated that MASHR and UDR significantly improved cross-population performance compared to the single-ancestry EN baseline (Figure 5). These results align with previous studies, which used MASHR to improve TWAS model generalizability across diverse populations and tissues^15,64^.

Furthermore, our findings complement recent improvements in East Asian brain tissue PWAS, which used an alternative multi-ancestry method called TransLasso, a transfer learning Lasso regularization technique. While TransLasso specifically leverages genetic knowledge from auxiliary datasets/populations (European and African) to improve estimations in a target population (East Asian)^65^, our application of MASHR and UDR provides a symmetric framework that jointly models the covariance structure across all ancestries simultaneously.

In *cis*-baseline testing scenarios, MASHR and UDR experienced a minor decline in predictive accuracy in ancestry-matched test populations compared to EN; however, across overlapping proteins, this impact was offset by the inclusion of *trans*-pQTLs and fine-mapping (Figure 6A). Notably, MASHR and UDR yielded a higher number of significantly predicted proteins across nearly all training and SNP-input conditions. However, performance remained consistently higher in EUR testing sets, reflecting the persistent sample size disparity between EUR (n = 1270) and AFR (n = 675) training cohorts, with an even larger disparity in the testing UKB data (EUR n = 49,009, AFR n = 1204).

Our analysis reveals that the optimal model architecture is likely protein-specific, driven by the unique regulatory mechanisms of each target. While the inclusion of *trans*-pQTLs improved the predictive performance of a specific subset of proteins, others exhibited a decline in accuracy due to the introduction of distal noise. EN and MASHR captured overlapping but distinct subsets of these *trans*-driven proteins, suggesting that different algorithmic approaches may be more adept at capturing specific distal signals. Similarly, we observed that while fine-mapping aims to isolate causal variants, it can inadvertently filter out informative SNPs, thereby reducing predictive power for certain proteins. This suggests that a one-size-fits-all approach to SNP selection is insufficient; instead, modeling efforts should differentiate between proteins driven by strong *cis*-acting variation and those governed by complex, distal regulatory networks. Given this, we provide an Ensemble model database of the best performing model for each protein that we recommend using in future PWAS applications.

Applying these models to PWAS across AoU and UKB cohorts demonstrated that the multivariate frameworks of UDR and MASHR identified more replicated associations than EN. Furthermore, multivariate discoveries spanned all ten tested phenotypes, whereas EN spanned only four. Crucially, AFR-trained multivariate models identified the most associations across all GWAS populations, outperforming even the larger EUR within-population analysis. This suggests that the high genetic diversity and smaller LD blocks characteristic of AFR ancestry capture more precise regulatory signals. However, because UKB is almost entirely European, our ability to validate discoveries in non-European contexts may be limited; future validation should include cohorts featuring higher proportions of diverse, multi-ancestry participants. Notably, 47% of replicated associations were absent from the GWAS Catalog^53^. While all three methods identified a novel downregulation of ALP1 associated with hyperlipidemia risk, potentially linked via lipid absorption regulation, other discoveries appear more novel. For instance, MASHR uniquely identified a positive association between CRLF1 and type 2 diabetes. Given CRLF1’s role in nervous system development, this suggests a potential mechanistic involvement in diabetic neuropathy, highlighting the need for further functional validation of these novel protein-trait associations.

Colocalization analysis of our replicated protein-trait associations tests if the most likely causal SNPs are shared between the GWAS and pQTLs, providing more evidence that SNPs are acting through protein level regulation. We found 11 protein-trait associations with a high likelihood of colocalization (PP.H4 > 0.8). Colocalization analysis identified shared causal variants between *cis*-pQTLs for APOE, SORT1, ANGPTL3, and CELSR2 and hyperlipidemia GWAS signals, likely acting through coordinated regulation of lipid metabolism. APOE and CELSR2 are directly implicated in low-density lipoprotein (LDL) cholesterol levels, ANGPTL3 in triglyceride levels, and SORT1 and CELSR2 reside within the CELSR2-PSRC1-SORT1 locus, which shows strong genetic association with remnant cholesterol^68,69^. In addition, IL4R and IL1RL1 colocalized with asthma GWAS signals, with IL-4 and IL-13 signaling coordinately upregulating IL1RL1 expression through STAT6 and IL-4 receptor-α, suggesting shared genetic regulation of chronic airway inflammation^70^. EDA2R colocalized with type 2 diabetes GWAS signals and represented a novel locus not previously catalogued in the GWAS catalog. EDA2R expression is elevated in obesity, insulin resistance, and type 2 diabetes, identifying it as a novel aging-associated receptor and potential therapeutic target for metabolic disease^71^. These results demonstrate that pQTL colocalization with disease GWAS signals can illuminate the biological mechanisms underlying protein-trait associations, revealing coordinated pathway-level regulation and supporting the utility of multi-ancestry pQTL mapping for identifying disease-relevant loci.

Despite these advancements, several limitations remain. First, the accuracy of Olink’s PEA methodology may be compromised by coding SNPs that alter antibody binding affinity, which is difficult to address due to proprietary antibody designs^70^. Second, while blood plasma is the non-invasive gold standard, it fails to capture localized, tissue-specific protein expression, and the ∼3000 proteins measured by Olink offer an incomplete snapshot of the total proteome.

Third, benchmarking fine-mapping models is constrained by the lack of known ground truth for causal variants, precision and recall were low across all fine-mapping models tested and thus the underlying statistical assumptions could be improved. Ultimately, improving fine-mapping and prediction model accuracy requires larger sample sizes, particularly in non-European ancestries. Public dataset disparities still bias predictive performance toward European populations, though our findings underscore the value of diverse cohorts in PWAS discovery^5,10,11,71,72^. Addressing these technical and ancestry imbalances is critical for refining future PWAS.

Overall, our study demonstrates the importance of ancestry diversity in the field of human genetics. Multi-ancestry fine-mapping leverages ancestry-specific LD structure to improve causal variant resolution beyond what single-ancestry approaches achieve, with African-ancestry individuals contributing disproportionately to this gain. By training models that leverage shared effect sizes across populations and incorporating distal genetic signals, we identified biological mechanisms underlying complex traits like hyperlipidemia, type 2 diabetes, and hypertension that were otherwise uncaptured. To advance precision medicine and improve health for all populations, future research must prioritize both the expansion of underrepresented population genomic datasets and the continued development of multi-ancestry methodologies that can accurately map the functional proteome.

### Data and code availability

Trans-Omics for Precision Medicine (TOPMed) Freeze 10b whole-genome sequence data (GRCh38) were obtained through the National Heart, Lung, and Blood Institute (NHLBI) TOPMed Program.

TOPMed Multi-Ethnic Study of Atherosclerosis (MESA), sponsored by the NHLBI, data are available via the dbGaP study NHLBI TOPMed: MESA and MESA Family AA-CAC (phs001416.v4.p1).

The 1000 Genomes Project (1KG) phase 3 GRCh37 VCF files (release date 2013/05/02) are available for download at https://ftp.1000genomes.ebi.ac.uk/vol1/ftp/release/20130502/.

This study used data from the All of Us Research Program’s Controlled Tier Dataset version 8, available to authorized users on the Researcher Workbench.

UK Biobank Pharma Proteomics Project Protein GWAS Summary Statistics (2023) are available for download at https://doi.org/10.7303/syn51364943.

Pan-UK Biobank GWAS Summary Statistics were downloaded from within the Pan-UK Biobank phenotype manifest (last updated 2023/03/01) and are available at https://pan.ukbb.broadinstitute.org/downloads/index.html.

Code used to conduct the analyses in this study are available via GitHub at https://github.com/ckrueger2/MESA_FM_PWAS_2026.

Our developed multi-ancestry TOPMed MESA proteome prediction models, fine-mapping output, and gene coding region boundaries used for analysis are available at https://doi.org/10.5281/zenodo.15483844.

## Data Availability

Trans-Omics for Precision Medicine (TOPMed) Freeze 10b whole-genome sequence data (GRCh38) were obtained through the National Heart, Lung, and Blood Institute (NHLBI) TOPMed Program. TOPMed Multi-Ethnic Study of Atherosclerosis (MESA), sponsored by the NHLBI, data are available via the dbGaP study NHLBI TOPMed: MESA and MESA Family AA-CAC (phs001416.v4.p1). The 1000 Genomes Project (1KG) phase 3 GRCh37 VCF files (release date 2013/05/02) are available for download at https://ftp.1000genomes.ebi.ac.uk/vol1/ftp/release/20130502/. This study used data from the All of Us Research Program's Controlled Tier Dataset version 8, available to authorized users on the Researcher Workbench. UK Biobank Pharma Proteomics Project Protein GWAS Summary Statistics (2023) are available for download at https://doi.org/10.7303/syn51364943. Pan-UK Biobank GWAS Summary Statistics were downloaded from within the Pan-UK Biobank phenotype manifest (last updated 2023/03/01) and are available at https://pan.ukbb.broadinstitute.org/downloads/index.html. Code used to conduct the analyses in this study are available via GitHub at https://github.com/ckrueger2/MESA_FM_PWAS_2026. Our developed multi-ancestry TOPMed MESA proteome prediction models, fine-mapping output, and gene coding region boundaries used for analysis are available at https://doi.org/10.5281/zenodo.15483844. 

https://doi.org/10.5281/zenodo.15483844

https://github.com/ckrueger2/MESA_FM_PWAS_2026

## Acknowledgments

This work is supported by the NIH National Human Genome Research Institute Academic Research Enhancement Award R15HG009569 (HEW). MESA and the MESA SHARe project are conducted and supported by the National Heart, Lung, and Blood Institute (NHLBI) in collaboration with MESA investigators. Support for MESA is provided by contracts 75N92025D00022, 75N92020D00001, HHSN268201500003I, N01-HC-95159, 75N92025D00026, 75N92020D00005, N01-HC-95160, 75N92020D00002, N01-HC-95161, 75N92025D00024, 75N92020D00003, N01-HC-95162, 75N92025D00027, 75N92020D00006, N01-HC-95163, 75N92025D00025, 75N92020D00004, N01-HC-95164, 75N92025D00028, 75N92020D00007, N01-HC-95165, N01-HC-95166, N01-HC-95167, N01-HC-95168, N01-HC-95169, UL1-TR-000040, UL1-TR-001079, UL1-TR-001420, UL1TR001881, and R01HL105756. The authors thank the MESA participants and the MESA investigators and staff for their valuable contributions. A full list of participating MESA investigators and institutions can be found at http://www.mesa-nhlbi.org/.

Molecular data for the Trans-Omics in Precision Medicine (TOPMed) program was supported by the National Heart, Lung, and Blood Institute (NHLBI). Genome sequencing for

“NHLBI TOPMed: MESA” (phs001416.v4.p1) was performed at the Broad Institute Genomics Platform (3U54HG003067-13S1, HHSN268201600034I). Proteomics for “NHLBI TOPMed:

MESA” (phs001416.v4.p1) was performed at Broad Institute and Beth Israel Proteomics Platform (HHSN268201600034I - 75N92020F00001, amendment P00004). Core support including centralized genomic read mapping and genotype calling, along with variant quality metrics and filtering were provided by the TOPMed Informatics Research Center (3R01HL-117626-02S1; contract HHSN268201800002I). Core support including phenotype harmonization, data management, sample-identity QC, and general program coordination were provided by the TOPMed Data Coordinating Center (R01HL-120393; U01HL-120393; contract HHSN268201800001I). We gratefully acknowledge the studies and participants who provided biological samples and data for TOPMed.

This research has been conducted using the UK Biobank Resource under application number 898835. The individual-level data analyses testing proteome prediction models were conducted on the Research Analysis Platform (https://ukbiobank.dnanexus.com). We would like to thank all the participants of UK Biobank for their vital contribution to the resource.

We gratefully acknowledge *All of Us* participants for their contributions, without whom this research would not have been possible. We also thank the National Institutes of Health’s *All of Us* Research Program for making available the GWAS summary statistics examined in this study.

## Declaration of generative AI and AI-assisted technologies in the writing process

During the preparation of this work the authors used Claude and Gemini in order to edit for grammar, style, and clarity. After using this tool/service, the authors reviewed and edited the content as needed and take full responsibility for the content of the publication.

