## Supplemental Notes and Figures for "Multi-ancestry modeling improves fine-mapping resolution, protein prediction, and discovery for proteome-wide association studies"

**Supplementary Note 1. TOPMed MESA Fine-Mapping.**

For all TOPMed MESA fine-mapping models, we restricted analysis to TOPMed MESA cis- and trans-pQTL windows passing FDR < 0.05. We fine-mapped cis- and trans-pQTL windows using TensorQTL Sum of Single Effects (SuSiE) implementation[1]. We applied the 'cis_susie' mode to fine-map cis-pQTL windows and 'trans_susie' mode to fine-map trans-pQTL windows. We filtered overlapping SNPs across trans-credible set windows to ensure that no SNP was included in more than one credible set and to exclude trans-SNPs overlapping with cis-pQTLs. After filtering out overlapping SNPs, only credible sets with PIPs that add up to at least 0.9 (coverage probability ≥ 0.9) were maintained in the final filtered output. Default parameters specified in the original TensorQTL documentation were used in both modes with the seed set to 123 to ensure reproducibility of the fine-mapping analyses.

We fine-mapped cis- and trans-pQTL windows using the SusieR package[2]. For trans-pQTL fine-mapping, we constructed and fine-mapped the full trans-window per chromosome that included all SNPs from our post-QC genotype files within 1 Mb before the minimum genomic trans-pSNP position and 1 Mb after the maximum genomic trans-pSNP position. Input for SusieR is the ancestry-specific LD, derived from individual-level genotype data, and the full phenotype variance matrix. The susie-fitted model was parameterized to closely mirror the TensorQTL SuSiE implementation: L = 10, null_weight = NULL, estimate_prior_method = 'EM'. We used the 'susie_get_cs' function with coverage = 0.95 and min_abs_corr = 0.5 to retrieve 95% credible sets.

We fine-mapped cis-pQTL windows using the Sum of Shared Effects (SuShiE) Python package[3] using 'sushie finemap' with default parameters and seed = 123. Input for SuShiE is individual-level genotype data stratified by ancestry (EUR, AFR, CHN, HIS) and the corresponding individual-level phenotype data in matrix format, producing a single fine-mapping output that jointly leverages all ancestries.

We fine-mapped cis-pQTL windows using the multi-ancestry extension of SuSiE, MultiSuSiE[4]. The MultiSuSiE model requires pre-computed LD matrices for each ancestry included in the analysis, which we calculated for the cis-windows using LDStore[5]. The model also requires summary statistics, which we extracted from the TensorQTL cis-nominal mode output by protein. Finally, the model input can only include SNPs that are present in every population that is analyzed, so we subset our summary statistics and LD matrices to these intersecting SNPs. Input data for MultiSuSiE is separate ancestry-specific LD panels and summary statistics, producing a single fine-mapping output that jointly leverages all ancestries. The parameters used to build the MultiSuSiE-fitted model were assigned to most closely represent the parameters used in TensorQTL's SuSiE implementation: rho = np.where(np.eye(4), 1, 0.75), L = 10, min_abs_corr = 0.5. We required credible sets to have a coverage greater than 0.95 and a minimum absolute correlation greater than 0.5 to reflect the purity filtering performed with TensorQTL.

We fine-mapped cis-pQTL windows using SuSiEx[6]. Like MultiSuSiE, SuSiEx requires summary statistics input. SuSiEx accepts PLINK files directly for calculating LD and does not require pre-computed LD matrices. Therefore, we used the same summary statistics used in MultiSuSiE and individual-level genotype data stratified by ancestry to produce a single fine-mapping output that jointly leverages all ancestries. We applied SuSiEx parameters '--keep-ambig = TRUE', '--mult-step = TRUE', and '--tol = 0.001' to most closely represent the parameters used in TensorQTL's SuSiE implementation.

**Supplementary Note 2. UKB Fine-Mapping.**

Pairwise LD calculations between cis-SNPs of each protein level phenotype are required for fine-mapping. We used the 1000 Genomes Project (1KG) phase 3 GRCh37 (release file date 2013/05/02) as a reference panel for LD matrix computation within each ancestry, as matched in-sample LD matrices were not available to us and ancestry-matched matrices for all five ancestral populations analyzed do not currently exist[7]. To assign ancestry to individuals within the reference panel, we used the integrated call samples panel (version 3, released 2013/05/02), which provides superpopulation ancestral classifications (EUR, AFR, AMR, EAS, SAS) for all 2504 reference individuals. We filtered the 1KG reference panel to only include SNPs, separated by Super Population to proceed with 4 populations: EUR (n = 503), AFR (n = 661), AMR (n = 347), EAS (n = 504), and built indexed .bgen files[8]. For each protein level phenotype, we constructed ancestry-stratified z-files by extracting cis-SNPs from the UKB summary statistics and matching variant identifiers to the corresponding 1KG reference panel .pvar file, retaining only SNPs present in both datasets.

Using the ancestry-specific protein level phenotype z-files and indexed .bgen files, we used LDStore[5] to write .bcor files containing pairwise SNP correlations for each phenotype within each ancestry (EUR, AFR, AMR, EAS). We then filtered each ancestry-protein phenotype combination to retain only SNPs present across all ancestries, with no additional variant-level filters applied, as required by downstream fine-mapping tools. We calculated ancestry weights as the proportion of each ancestry's sample size relative to the total UKB population (EUR: 33,187, AFR: 931, AMR: 97, EAS: 262, total: 34,477), and we computed the final META LD matrix as the weighted sum of the ancestry-specific filtered LD matrices. We filtered ancestry-specific protein level summary statistics to include only SNPs present across all four UKB ancestries and the 1KG LD reference panels.

For all UKB fine-mapping models, we restricted analysis to cis-windows with respective TOPMed MESA cis-pQTL windows passing FDR < 0.05 that also produced at least 1 credible set in TensorQTL cis-SuSiE fine-mapping. We fine-mapped cis-pQTL windows using SusieR[2], SuShiE[3], MultiSuSiE[4], and SusieX[6], applying the weighted 1KG META LD matrix and UKB summary statistics for each protein-level phenotype.

For UKB SusieR fine-mapping, we applied the 'susie_rss' model using the same parameters as in TOPMed MESA fine-mapping, except for the sample size parameter (n = 34,477).

For UKB SuShiE fine-mapping, we applied the same parameters as in TOPMed MESA SuShiE fine-mapping, with the addition of '--ld-adjust 0.02', a ridge regularization approach that adds a small number to the diagonal of the LD matrix to ensure LD matrices are positive semi-definite, which is required by SuShiE[9].

MultiSuSiE requires summary statistics and pre-computed pairwise LD matrices, therefore we used the same parameters for UKB fine-mapping as TOPMed MESA fine-mapping.

For UKB SuSiEx fine-mapping, when individual-level genotype data is not used, SuSiEx requires ancestry-specific '_frq.frq', '.ld.bin', and '_ref.bim' files as its LD input. We used Python to reformat the ancestry-specific 1KG LD matrices into the required format for each protein-ancestry combination. We used identical parameters to our TOPMed MESA SuSiEx fine-mapping.

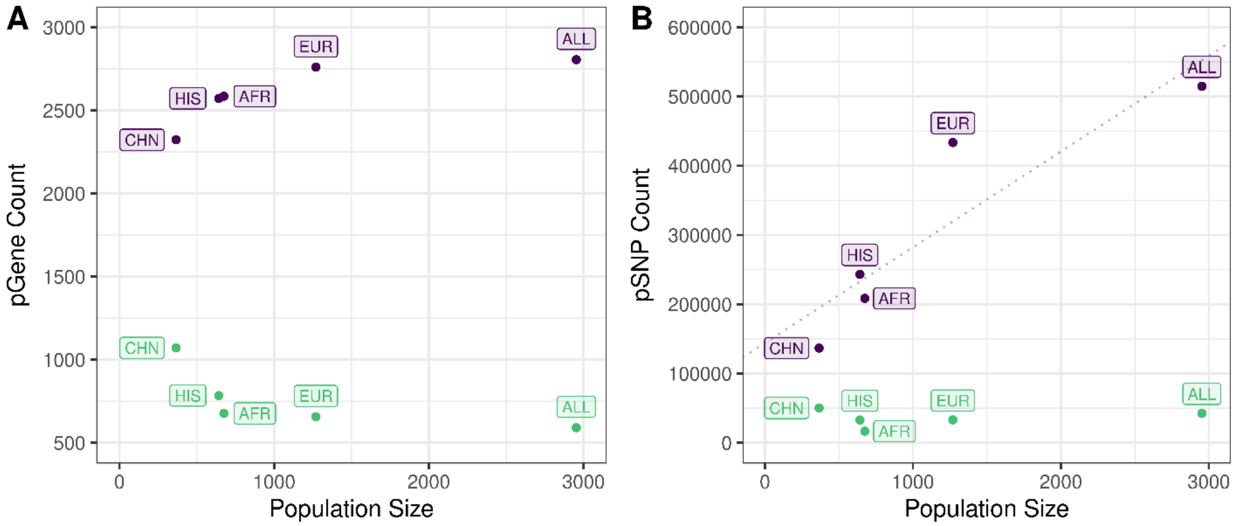


**Figure S1. Visualization of opposing discovery patterns between cis-pQTLs and trans-pQTLs.** (A) Number of proteins with at least one FDR-significant cis- (purple) or trans-pSNP (green) identified by TensorQTL pQTL mapping, plotted against TOPMed MESA population sample size. (B) Relationship between the cis- (purple) and trans-pSNP (green) counts across proteins, plotted against TOPMed MESA population sample size. Linear regression for cis-pSNPs is displayed by purple dashed line (slope = 138.191, $R^2$ = 0.816, p = 0.035), and was the only significant correlation between pGene and pSNP to population size analyses.

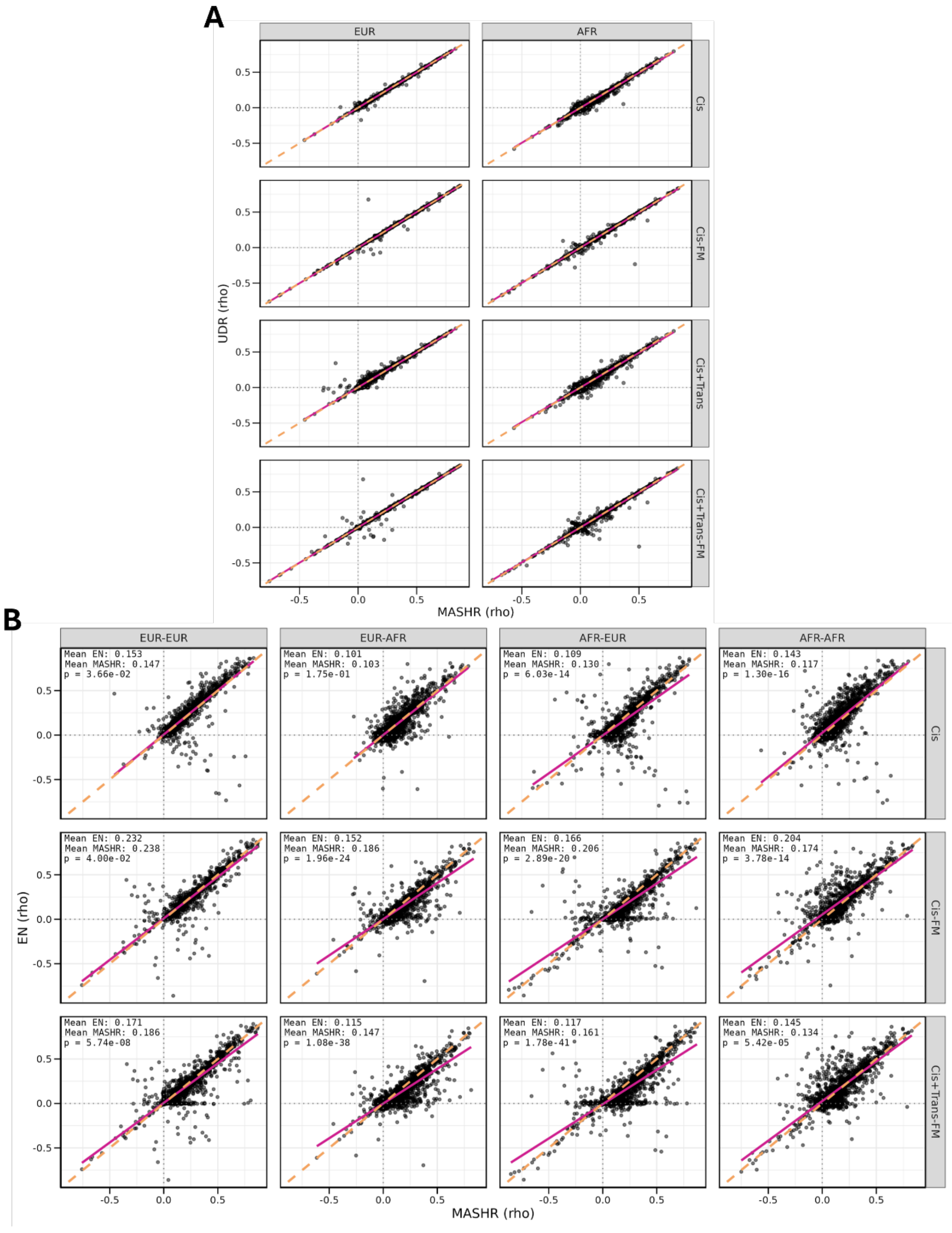


**Figure S2.** (A) Pairwise scatterplots comparing the within-population (TOPMed MESA EUR-UKB EUR, TOPMed MESA AFR-UKB AFR) predictive accuracy (ρ) of MASHR (x-axis) vs UDR (y-axis) across SNP input types. Each point represents an individual protein. The number of proteins varies by input type based on the availability of significant fine-mapped or trans variants: n = 2703 for Cis comparisons, n = 1212 for Cis fine-mapped comparisons, n = 2850 for Cis+Trans comparisons, and n = 1935 for Cis+Trans fine-mapped comparisons. The pink line represents the regression lines between conditions. The orange dash line represents the identity line (y = x). (B) Pairwise scatterplots comparing the within-population (EUR-EUR, AFR-AFR) and cross-population (EUR-AFR, AFR-EUR) predictive accuracy (ρ) of MASHR (x-axis) vs EN (y-axis) across SNP input types. Each point represents an individual protein. The number of proteins varies by input type based on the availability of significant fine-mapped or trans variants: n = 2339 for Cis comparisons, n = 1199 for Cis fine-mapped comparisons, and n = 1883 for Cis+Trans fine-mapped comparisons. The pink line represents the regression lines between conditions. The orange dash line represents the identity line (y = x).

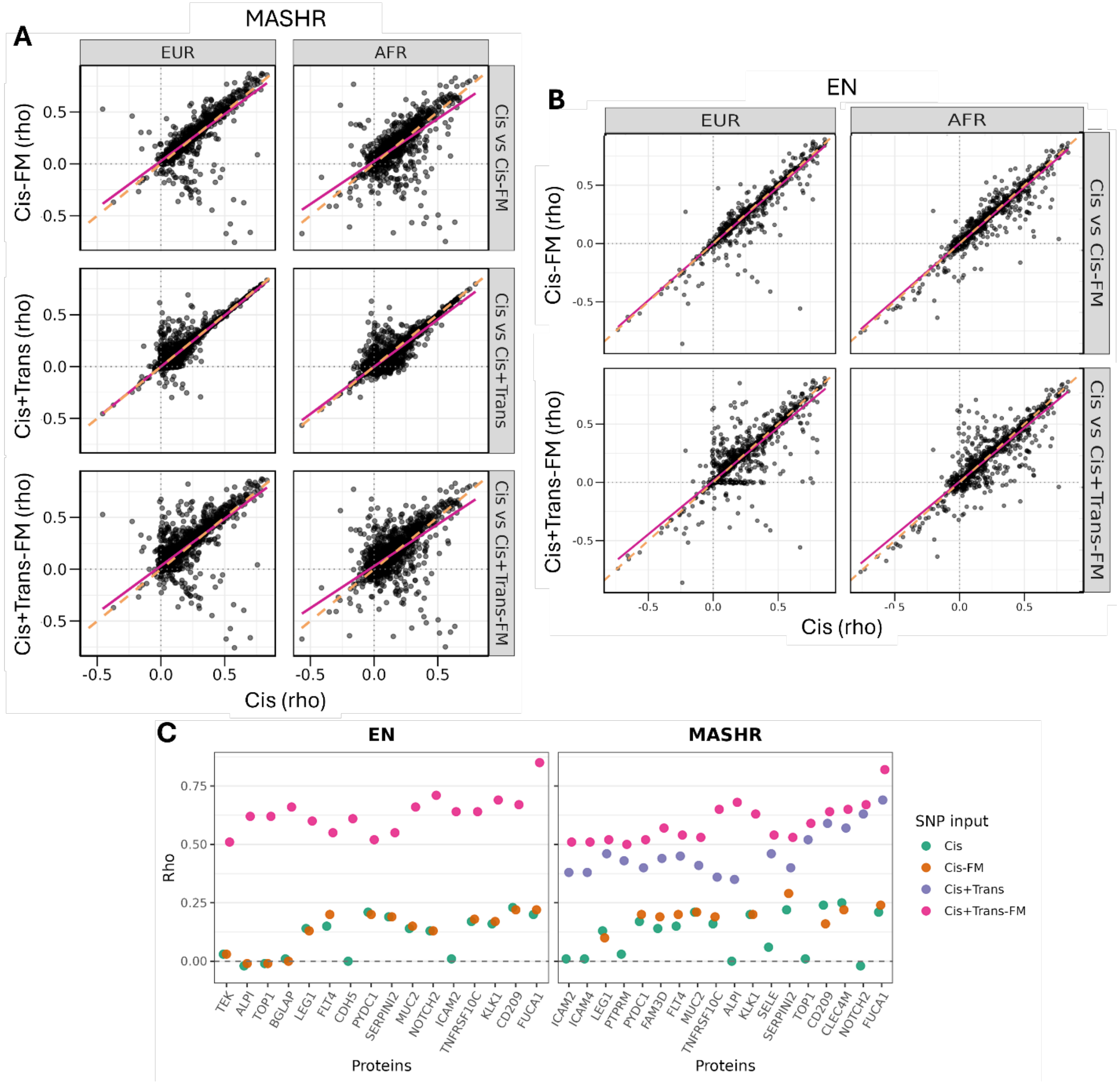


**Figure S3.** (A) Pairwise scatterplots comparing the within-population (EUR-EUR, AFR-AFR) predictive accuracy (ρ) of the MASHR cis-baseline (x-axis) against expanded SNP input strategies (y-axis: Cis-FM, Cis+Trans, and Cis+Trans-FM, respectively). Each point represents an individual protein. The number of proteins varies by input type based on the availability of significant fine-mapped or trans variants: n = 1212 for cis-FM comparisons (top), n = 2862 for cis+trans comparisons (middle), and n = 1946 for cis+trans-FM comparisons (bottom). The pink line represents the regression lines between conditions. The orange dash line represents the identity line (y = x). (B) Pairwise scatterplots comparing the within-population (EUR-EUR, AFR-AFR) predictive accuracy (ρ) of the EN cis-baseline (x-axis) against expanded SNP input strategies (y-axis: Cis-FM and Cis+Trans-FM, respectively). Each point represents an individual protein. The number of proteins varies by input type based on the availability of significant fine-mapped or trans variants: n = 1762 for cis-FM EUR, n = 1640 for cis-FM AFR, n = 1826 for Cis+Trans-FM EUR, n = 1693 for Cis+Trans-FM AFR.The pink line represents the regression lines between conditions. The orange dash line represents the identity line (y = x). (C) Protein subset with cis and cis-FM $\rho < 0.25$ and cis+trans and cis+trans-FM $\rho > 0.5$ in both EUR and AFR plots between EN and MASHR, showing predictive accuracy (rho, ρ) changes on the y-axis and proteins on the x-axis.

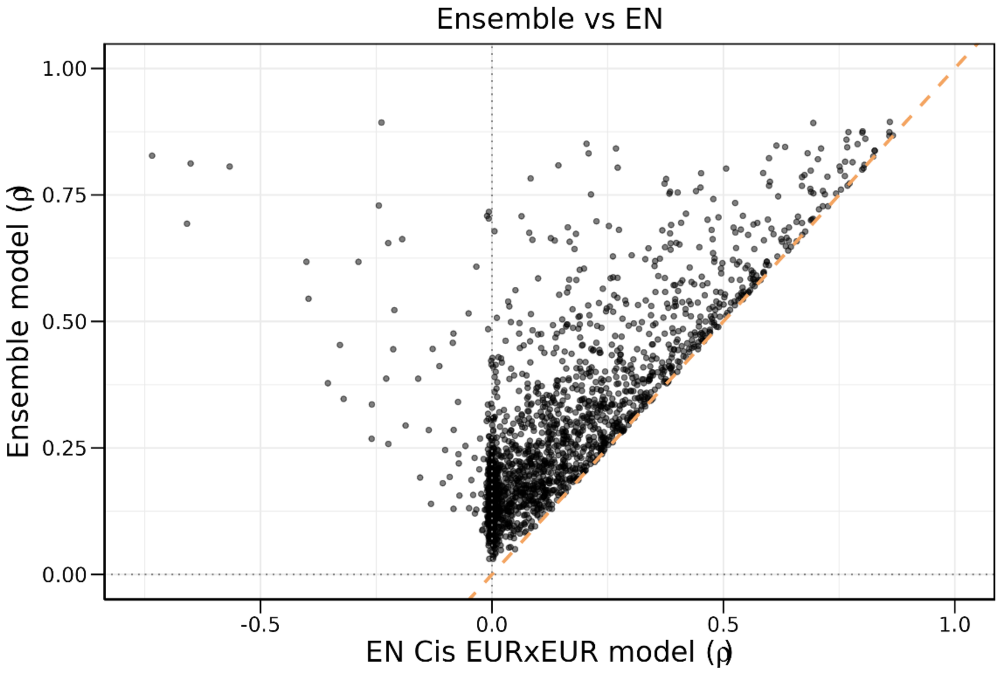


**Figure S4.** Pairwise scatterplots comparing the Ensemble model vs EN Cis EUR-EUR model predictive accuracy (ρ). Each point represents an individual protein, with 1934 overlapping protein models compared. The orange dash line represents the identity line (y = x).

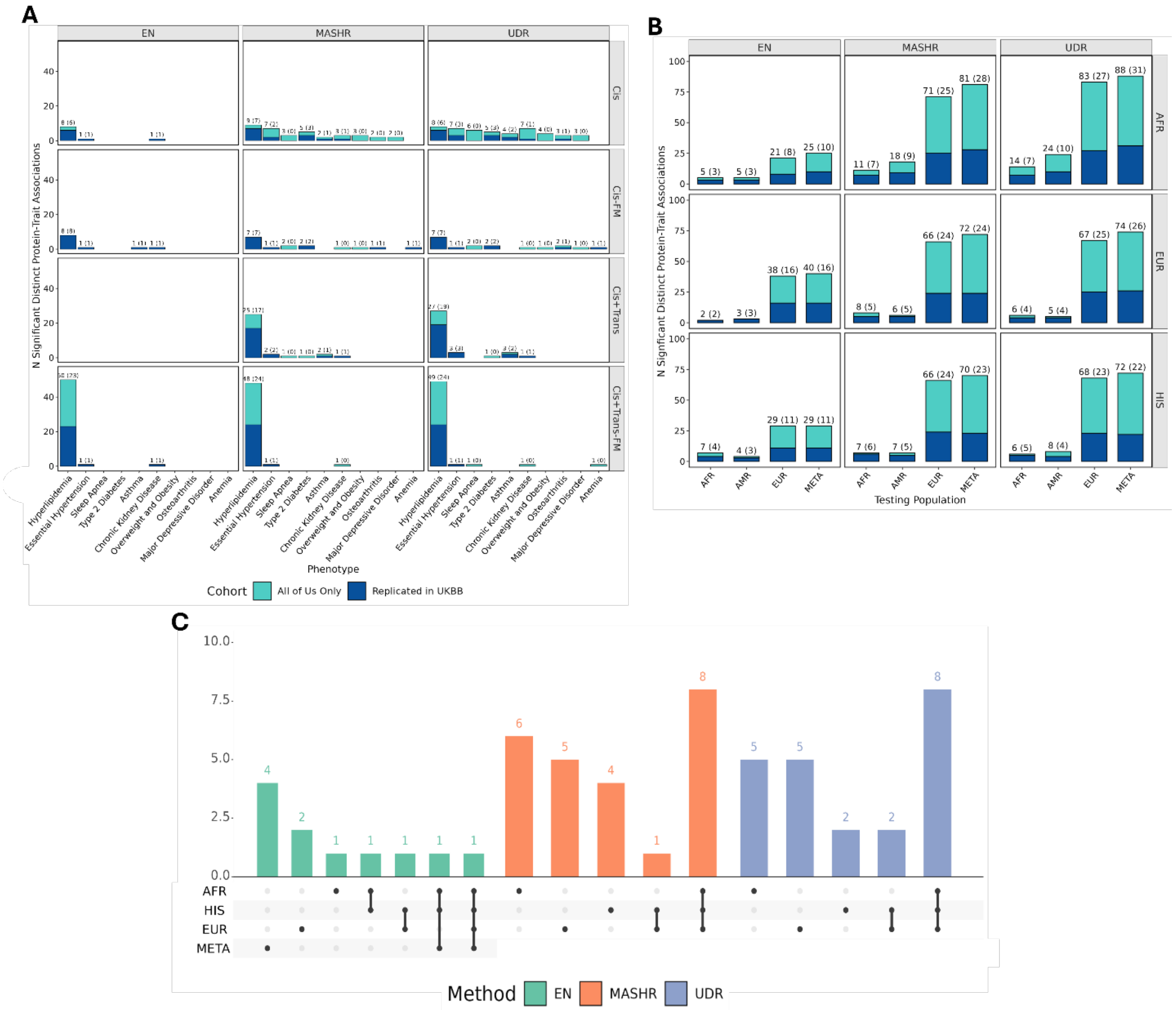


**Figure S5.** (A) Significant distinct associations between phenotypes, methods, and SNP types. Total AoU discoveries are indicated by the height of the stacked bar, with the subset of UKB-validated associations highlighted in navy. (B) Significant distinct associations with hyperlipidemia are shown for EN, MASHR, and UDR across African American (AFR), European (EUR), and Hispanic/Latino (HIS) training models and African (AFR), European (EUR), Admixed American (AMR), and META AoU and UKB testing populations. Total AoU discoveries are indicated by height of the stacked bar, with the subset of UKB-validated associations highlighted in navy. (C) Upset plot comparing significant distinct protein-trait associations within each machine learning method across training populations.
